# Exome Sequencing Identifies *POPDC2* as a Candidate Gene for Familial Congenital Junctional Ectopic Tachycardia

**DOI:** 10.64898/2026.05.12.26353039

**Authors:** Benjamin M. Helm, Alexander H. Swan, Susanne Rinné, Mark Pfuhl, Emilia De Martino, Adam C. Kean, Niels Decher, Thomas Brand

## Abstract

**Background:** Congenital junctional ectopic tachycardia (cJET) is a rare, potentially life-threatening arrhythmia suspicious for a genetic basis, yet its molecular underpinnings remain incompletely defined. The *POPDC2* gene, involved in cardiac pacemaking and membrane trafficking of interacting ion channels, has not previously been conclusively linked to human tachyarrhythmias. This study investigates a novel *POPDC2* variant (p.Leu245Pro) identified in a family with autosomal dominant cJET.

**Methods:** Exome sequencing was performed to identify co-segregating variants in the affected family. Functional analysis of the *POPDC2* p.Leu245Pro variant was conducted by molecular dynamics (MD) simulations, a membrane targeting assay, and a bimolecular fluorescence complementation assay. Additionally, the impact of the variant on Nav1.5 and TREK-1 currents was characterized in *Xenopus* oocytes.

**Results:** The p.Leu245Pro POPDC2 variant showed a destabilization of the POPDC1-POPDC2 dimer interface, resulting in impaired heterodimer formation and membrane localization. Electrophysiological studies in *Xenopus* oocytes demonstrated that the mutant protein significantly affected Nav1.5 and TREK-1 currents. These findings support a functional impact of the *POPDC2* p.Leu245Pro variant relevant to cardiac conduction.

**Conclusions:** Our results provide the first functional evidence implicating *POPDC2* in cJET and support its role as a novel candidate gene in tachyarrhythmic disease. This study enhances the understanding of genetic contributions to cJET and suggests further investigation of *POPDC2* in other forms of supraventricular tachyarrhythmias.

## INTRODUCTION

An automatic tachycardia originating in the atrioventricular (AV) node or His bundle is termed junctional ectopic tachycardia (JET). JET in the pediatric population is commonly divided into three varieties: (1) The congenital (cJET)—also known as His bundle tachycardia—that is a rare tachyarrhythmia in infants and can also be diagnosed prenatally; (2) the possibly acquired automatic junctional tachycardia that is diagnosed in children >6 months, adolescents, and adults; and (3) the much more commonly recognized post-operative JET that is often encountered following cardiac surgery ^1^. cJET is very rare, with an international multicenter review describing only 44 patients over 40 years from the 1960s to the 2000s ^2^. It is generally defined by an onset *in utero* or under 6 months of life and is often incessant, leading to significant cardiac dysfunction, while surgical JET has been attributed to inflammation or perturbation of the AV junction that may accompany surgical manipulation. Importantly, while focal JET is thought to be secondary to increased automaticity similar to atrial ectopic tachycardia, the specific causes of cJET and post-operative JET remain unclear. Accordingly, optimal treatment has remained equally elusive, with primary strategies involving amiodarone vs. procainamide, though some success with amiodarone and adjunctive ivabradine, as well as sotalol, is showing promise ^3,4^. Enhanced automaticity is the primary suggested pathophysiological mechanism proposed for cJET, as this has been shown to be the cause of acquired focal JET ^5^.

There are a few genetic associations described for post-operative JET and cJET, and there may be familial predispositions in up to 50% of cases ^6–9^, however, due to its rarity, limited genetic investigations are using contemporary genomic sequencing methods ^1,10^. To date, the only putative monogenic cause of familial/autosomal dominant JET is the *TNNI3K* gene encoding Troponin I-interacting kinase, a member of the MAP kinase kinase kinase (MAPKKK) family of protein kinases, and variants with reduced kinase activity have been linked to cJET ^11^. However, these reports included cases with complex presentations such as variable AV block (AVB), atrial tachyarrhythmias, and dilated cardiomyopathy ^8,12^. Isolated cases of JET associated with viral myocarditis have also been reported ^13^. Work in mice has identified *JPH2* encoding Junctophilin-2, a structural protein that establishes cardiac dyads as candidate gene for causing cJET ^14,15^. However, *JPH2* variants have not yet been genetically linked to cJET.

Family-based study designs should be fruitful for identifying putative genetic causes of cJET. However, this approach is of limited use for cJET, owing to the ultra-rarity of familial cJET and the small family sample sizes. While population-scale biobanks with genomic data from unrelated cases and controls are popular for disease-gene identification, family-based studies can be more powerful—especially for detecting rare variants co-segregating with rare phenotypes^16–19^. These study designs, combined with genomic sequencing and rare-variant association analyses, could be advantageous for investigating genetic causes of cJET.

In this study, we completed exome sequencing in a family of four affected by cJET diagnosed/treated at one of our institutions and investigated potential candidate gene variants. Following prioritization, a novel *POPDC2* (p.Leu245Pro) variant was identified, which might be causally associated with the cJET phenotype. *POPDC2* is a member of the Popeye domain containing (POPDC) gene family encoding transmembrane proteins, which function as cAMP effector proteins^20^. *POPDC2* is expressed at high levels in cardiac myocytes and shows a pronounced expression in the cardiac conduction system^21^. It functions in cardiac pacemaking and AV conduction^21^, and regulates membrane trafficking of the K_2P_ potassium channel *KCNK2*/TREK1^21^ and the sodium channel Nav1.5^22^. These properties lead us to prioritize the *POPDC2* variant for functional studies. Molecular dynamics (MD) simulation of the POPDC1-POPDC2 protein complex revealed a distortion and destabilization of the dimer interface in the POPDC2 p.Leu245Pro mutant. Using a recently established membrane targeting assay ^23^, we observed abnormal membrane localization of POPDC2 p.Leu245Pro. Moreover, a bimolecular fluorescence complementation (BiFC) assay ^23^ demonstrated aberrant complex formation of the mutant POPDC2 protein with POPDC1. Measurements of Nav1.5 and TREK-1 currents in *Xenopus* oocytes ^21,22^ after co-expression of wild-type and mutant POPDC2 also provided evidence for altered protein function of the POPDC2 p.Leu245Pro variant. We thereby provide strong supportive functional evidence of the contribution of the mutant *POPDC2* allele to familial cJET and suggest future investigation of *POPDC2* in supraventricular tachyarrhythmias. In addition, this study helps to improve our knowledge of possible genetic associations with cJET, and our results should additionally possibly also help to improve our understanding of other forms of JET—including post-operative/iatrogenic JET.

## METHODS

### Ethics

The family-based case study was approved by the Indiana University/Indiana University Health Institutional Review Board (IRB#: 18113646211) with a waiver of consent and exempt from review. No identifying information is shared, and no studies of human subjects were performed. Experiments using *Xenopus laevis* toads were approved by the ethics committee of the Regierungspräsidium Giessen (protocol code: V5419c2015h01MR20/28 Nr. V16/2022).

### Case Ascertainment and Clinical Evaluations

Members of the affected family were seen for clinical evaluation and medical management in the pediatric heart rhythm clinic at Riley Hospital for Children at Indiana University Health. All affected relatives had been initially diagnosed and treated at our center, and the affected father continued his adult cardiology care outside of our center. All patients were evaluated by board-certified pediatric cardiologists/electrophysiologists.

### Clinical Exome Sequencing

We completed genetic testing as part of the clinical evaluation. Genetic testing consisted of family-based exome sequencing, i.e., quad/tetrad exome sequencing, including the affected relatives across two generations in this family. Notably, we leveraged the family structure to prioritize testing in all affected relatives, including two full siblings, a paternal half-sibling, and the father of the proband; due to family structure, we were unable to test clearly unaffected relatives. Clinical-grade exome sequencing was performed by a commercial clinical laboratory (GeneDx, LLC, Gaithersburg, MD). Methods for exome sequencing are detailed and modified from the clinical report. Genomic DNA was extracted directly from the submitted specimens (buccal swab). The DNA was enriched for the complete coding regions and splice site junctions for most genes of the human genome using a proprietary capture system developed by GeneDx for next-generation sequencing with copy-number variant (CNV) calling (NGS-CNV). The enriched targets were simultaneously sequenced with paired-end reads on an Illumina platform. Bi-directional sequence reads were assembled and aligned to reference sequences based on NCBI RefSeq transcripts and human genome build GRCh37/UCSC hg19. Using a custom-developed analysis tool at GeneDx (XomeAnalyzer), data were filtered and analyzed to identify sequence variants and most deletions and duplications involving three or more coding exons ^24^. Smaller deletions or duplications may not be reliably identified. Reported variants were confirmed, if necessary, by an appropriate orthogonal method in the proband and, if submitted, in selected relatives. Sequence variants are reported according to the Human Genome Variation Society (HGVS) guidelines. CNVs are reported based on the probe coordinates, the coordinates of the exons involved, or precise breakpoints when known. Reportable variants include pathogenic variants and likely pathogenic variants. Variants of uncertain significance, likely benign and benign variants, if present, are not routinely reported. Available evidence for variant classification may change over time and variant(s) may be reclassified according to the ACMG/AMP Standards and Guidelines ^25^. CNV detection was performed using the data. Mean depth of coverage was 98x with quality threshold of 98.7%.

### Independent Variant Review and Novel Candidate Gene Prioritization

The family provided written consent to have data released from the laboratory to the clinical team (BMH) in the form of exome sequencing alignment files (CRAM) and variant annotation files (VCF) for each of the submitted samples/tested relatives. Data were stored in a secure, HIPAA-compliant location. The VCF files were sorted, compressed, and merged into a final family VCF file using Samtools and BCFtools ^26^. Individual information was preserved and referenced in a family (PED) file for downstream analyses, and affected status was defined for each relative in the PED file in binary fashion. We used Exomiser to prioritize candidate gene variants in the VCF files potentially associated with the patient/family phenotype ^27,28^. Individual information was preserved and referenced in a family (PED) file for downstream analyses, and affected status was defined for each relative in the PED file in a binary fashion. Briefly, Exomiser uses multiple algorithms to prioritize gene discovery and variant-phenotype associations based on protein interaction networks, clinical relevance, cross-species phenotype comparisons, population prevalence, predicted pathogenicity/deleteriousness, and inheritance pattern/family segregation when available ^27–29^. Additional studies have reported on the benefits of the Exomiser *PhenIX*/*hiPHIVE* algorithms for successfully prioritizing causative gene variants, especially for phenotype-driven novel gene discovery ^27,30^. For our Exomiser analysis, we used the standard thresholds for variant quality, filtering, and prioritization for presumed rare/ultra-rare autosomal dominant diseases and mode of inheritance as recommended by the authors of Exomiser ^29,31^. We used the Human Phenotype Ontology (HPO) term for junctional ectopic tachycardia (HP:0011716) as the only phenotype descriptor in our analysis. Default settings for minor allele frequency (MAF) thresholds were used; more specifically, we used a MAF threshold of 0.1% for autosomal dominant inheritance for consideration of candidate gene variants (cross-referenced against ExAC, gnomAD, ESP, and 1000 Genomes population databases as standard for Exomiser). We also filtered variants for autosomal recessive, X-linked, and mitochondrial inheritance patterns; however, the family phenotype is strongly suggestive of an autosomal dominant pattern. Therefore, we only prioritized variants that were co-segregating in all affected relatives in the family. We then qualitatively assessed the composite Exomiser score assigned to each variant, and we classified variants with composite scores ≥0.80 as strong candidates; otherwise, we also reviewed variants with composite scores ≥0.75 as moderate candidates. We also included Rare Exome Variant Ensemble Learner (REVEL) and Combined Annotation-Dependent Depletion (CADD, v1.6) scores to compare predicted deleteriousness of the top 22 prioritized missense variants when able ^32,33^. We prioritized variants with CADD ≥25 and REVEL ≥0.75 as supporting evidence for deleteriousness/pathogenicity ^32,34^. The latter score has been shown to result in higher specificity, but slightly reduced sensitivity as compared to REVEL threshold of ≥0.50.

### Cell culture

HEK293 (DSMZ) cells were cultured in Dulbecco’s modified Eagle’s medium (Sigma Aldrich) supplemented with 10% (v/v) fetal bovine serum (Merck). Cells were transiently transfected using calcium phosphate (Promega), followed by a 48-h post-transfection incubation period before analysis.

### Cloning procedures

Full-length human *POPDC1* and *POPDC2* cDNAs were inserted into the pECFP-N1 and pEYFP-N1 plasmids to append C-terminal ECFP/EYFP tags. The *POPDC2* p.Leu245Pro mutant sequence was produced by site-directed mutagenesis using the Q5 site-directed mutagenesis kit (NEB). The oligonucleotide primer sequences used for the mutagenesis are: 5’-TCGGAGAAGCCCTACACTCTC-3’ (forward primer) and 5’-GATGTCATATCCCA GCAG-3’ (reverse primer). The constructs used in bimolecular fluorescence complementation (BiFC) experiments were created through the ligation of full-length wild-type and mutant POPDC cDNAs into the pECFP-N1 or pEYFP-N1 plasmids, respectively (kindly provided by Dr. Carmen Dessauer, University of Texas, Houston), using NotI and BamHI restriction sites. For injection into *Xenopus* oocytes, the POPDC2 p.Leu245Pro mutant sequence was cloned into the oocyte expression vector pSP64.

### Co-expression of POPDC isoforms in HEK293 cells

HEK293 cells were transiently transfected with wild-type POPDC1 and either wild-type POPDC2 or POPDC2 p.Leu245Pro constructs, which contained C-terminal enhanced cyan fluorescent protein (ECFP) or enhanced yellow fluorescent protein (EYFP) tags, respectively. All wild-type and mutant samples were transfected, processed and imaged concurrently across ≥2 experiments 24 hours before imaging, cells were seeded into poly-L-lysine-coated 8-well microscope slides with a D263 M Schott glass, No. 1.5H, 170 μm ± 5 μm glass coverslip base (Ibidi). On the day of imaging, the plasma membrane of the cells was marked through incubation for 12 min at 37 °C with culture medium supplemented with 0.5% (v/v) CellBrite Red solution (Biotium), which contains 1,1′- dioctadecyl-3,3,3′,3′-tetramethylindo-dicarbocyanine (DiD). Following washes with PBS, cells were fixed with PFA. Nuclei were stained with Hoechst-33342, and cells were immersed in PBS for imaging. An LSM 780 Axio Observer inverted confocal laser scanning microscope (Zeiss), with a plan-apochromat 63X/1.40 oil objective (Zeiss), was used to image the cells. The Hoechst-33342, POPDC1-ECFP, POPDC2-EYFP and DiD fluorophores were detected using the following excitation and emission settings, respectively: 405 nm/410–452 nm, 458 nm/463–516 nm, 514 nm/519–621 nm and 633 nm/636–735 nm.

### Image analysis of transfected cells

FIJI was used to analyze all images ^35^. Outlines of the plasma membrane of the HEK293 cells were constructed, using the DiD channel as a guide. A fixed radial dilation of 0.8 µm was applied to the plasma membrane outline to generate a membrane region of interest (ROI). This width, although greater than the ultrastructural thickness of the plasma membrane, was empirically determined to optimally capture the DiD-labelled membrane signal while accommodating contour irregularities and fluorescence spread. This was applied uniformly across all images. The cytoplasmic ROI was defined as the area between the inner boundary of the membrane ROI and the nuclear boundary, which was automatically segmented using the Hoechst-33342 channel. Nuclear regions were excluded from analysis. After subtraction of background fluorescence, the average intensity of ECFP (POPDC1) and EYFP (POPDC2) fluorescence from the plasma membrane and cytoplasmic compartments of each cell was determined. The plasma membrane-to-cytoplasm fluorescence ratio was calculated individually for POPDC1-ECFP and POPDC2-EYFP or POPDC2-EYFP p.Leu245Pro within each cell.

### Bimolecular fluorescence complementation

Constructs of full-length wild-type POPDC1 and wild-type POPDC2 or the p.Leu245Pro mutant, with respective C-terminal VC155 or VN155 split Venus domains, were co-expressed in HEK293 cells_across ≥2 experiments. Transfection with single POPDC1- VN155 and POPDC2-VN155 constructs was used as a negative control to assess background fluorescence. Transfection with pmRFP-N1 alongside POPDC constructs was performed to provide an internal control for transfection efficiency to which the Venus signal could be normalized. 48 hours after transfection, the cells were fixed using 4% (w/v) PFA and nuclei stained using Hoechst-33342. The cells were imaged using an Axio Observer inverted confocal laser scanning microscope (Zeiss) using a 10X objective (Zeiss). The signals of Hoechst-33342, reformed Venus and mRFP fluorophores were measured using the following excitation/emission settings, respectively: 405 nm/410–503 nm, 514 nm/516–587 nm, and 543 nm/582–754 nm. FIJI was used to analyze the cells. A mask was created that contained all cells which possessed levels of mRFP fluorescence that were above background, thus excluding non-transfected cells. The nuclei of cells were defined using the Hoechst-33342 channel and were excluded from the analysis. The median Venus intensity across all transfected cells within each image was then determined and normalized to the median mRFP fluorescence. The average normalized Venus fluorescence across all images from each group was then found.

### Sequence alignments and structural models of POPDC proteins

For the sequence alignment of the αC-helix, amino acid sequences of homo and paralogs were retrieved from the UniPROT database (www.uniprot.org): POPDC1: *Homo sapiens* (Q8NE79, residues: 243-267), *Mus musculus* (Q9ES83, residues: 243-267), *Gallus gallus* (Q9DG23, residues: 240-264), *Danio rerio* (Q5PQZ7, residues: 233-257), invertebrate POPDC proteins: *Branchiostoma lanceolatum* (A0A8J9YK61, residues: 255-279), *Drosophila melanogaster* (Q9VRE2, residues: 297-321), *Hydra vulgaris* (T2M9R6, residues: 221-245), POPDC2: *Homo sapiens* (Q9HBU9, residues: 227-251), *Mus musculus* (Q9ES82, residues: 227-251), *Gallus gallus* (Q6TRX0, residues: 227-251), *Danio rerio* (Q6JWV8, residues: 229-253), POPDC3: *Homo sapiens* (Q9HBV1, residues: 229-253), *Mus musculus* (Q9ES81, residues: 225-249), *Gallus gallus* (Q9DG25, residues: 227-251), *Danio rerio* (Q6JWW1, residues: 228-252).

### Popeye Domain Interaction Modelling

The model of the heterodimer of the Popeye domains of human POPDC1 and POPDC2 was generated using the AlphaFold3 ^36^ server using the UniPROT sequences of POPDC1 (Q8NE79) and POPDC2 (Q9HBU9). Structure images were produced with ChimeraX ^37^ and Yasara ^38^.

### MD simulations

Using Yasara ^38^, leucine 245 of POPDC2 was substituted by proline (POPDC2 p.Leu245Pro) or aspartate (POPDC2 p.Leu245Asp) in the AlphaFold3 model of the POPDC1-POPDC2 heterodimer using the ‘swap’ function, after which hydrogens were added using the ‘Clean’ function. The modelled WT and mutant heterodimers were then subjected to a steepest descent and then simulated annealing energy minimization to relax the structures and to remove unfavorable interactions. MD simulations were carried out for heterodimers of POPDC1 and wild-type POPDC2, POPDC2 p.Leu245Pro or POPDC2 p.Leu245Asp, respectively for 1 µs. The MD simulations were run with Yasara version 23.5.19^38^. Prior to the MD simulation, the hydrogen bonding network was optimized ^39^ to increase the solute stability, and a pKa prediction to fine-tune the protonation states of protein residues at the chosen pH of 7.0. The protein was placed in a dodecahedral cell with a diameter 5 Å larger than the largest dimension of the protein to avoid boundary effects of the rotating protein. The cell parameters are 75 x 75 x 75 Å with angles 60, 90 and 90° and a volume of 628661 Å^3^. It was filled with 8377 water molecules to which NaCl ions were added with a physiological concentration of 0.9%, with an excess of either Na^+^ (25) or Cl^-^ (23) to neutralize the cell. The simulation trajectories were run using the AMBER14 force field ^40^ for the solute and TIP3P for water. The cut-off was 8.0 Å for Van der Waals forces (the default used by AMBER ^41^ while electrostatic interactions were fully calculated with the Particle Mesh Ewald algorithm ^42^. The equations of motion were integrated with a multiple timestep of 2.5 fs for bonded and 5.0 fs for non-bonded interactions at a temperature of 298K and a pressure of 1 atm (NPT ensemble) using algorithms described in detail previously ^38^. Snapshots of the trajectory were stored in intervals of 100ps for subsequent analysis. Analysis of the MD trajectories was carried out using standard Yanaconda macros provided with the Yasara distribution or home written.

### Isolation of *Xenopus* laevis oocytes, cRNA synthesis and injection

Oocytes were isolated from anesthetized *Xenopus laevis* frogs and incubated in OR2 solution containing in mM: 82.5 NaCl, 2 KCl, 1 MgCl_2_, 5 HEPES; pH 7.5 with NaOH, supplemented with collagenase (1.5 mg/ml) (Nordmark) to remove residual connective tissue. Subsequently, oocytes were stored in ND96 solution containing in mM: 96 NaCl, 2 KCl, 1.8 CaCl_2_, 1 MgCl_2_, 5 HEPES; pH 7.4 with NaOH, supplemented with Na-pyruvate (275 mg/l), theophylline (90 mg/l) and gentamicin (50 mg/l) at 18 °C. Nav1.5 cDNA was linearized with XbaI and cRNA synthesis was done with the mMESSAGE mMACHINE™ SP6 Kit (ambion). TREK-1 and POPDC2 cDNAs were linearised with NheI, and cRNA was synthesized with the HiScribe® T7 ARCA mRNA Kit (New England Biolabs). Quality was tested using agarose gel electrophoresis and cRNAs were quantified by a spectrophotometer (NanoDrop, Thermo Fisher Scientific). Stage IV and V oocytes were each injected with 50 nl of cRNA.

### Two-electrode voltage-clamp recordings

Voltage-clamp recordings were performed with a TurboTEC 10CD (npi) amplifier and a Digidata 1200 Series (Axon Instruments) as A/D converter at room temperature (20 – 22 °C). Micropipettes were pulled from borosilicate glass capillaries GB 150TF-8P (Science Products) with a DMZ-Universal Puller (Zeitz). The resistance of the recording pipettes was 0.3-0.6 MΩ when pipettes were filled with 3 M KCl solution. The recording solution (ND96) contained in mM: NaCl 96, KCl 2, CaCl_2_ 1.8, MgCl2 1, HEPES 5 (pH 7.5). Data were acquired with Clampex 10 (Molecular Devices) and analyzed with Clampfit 10 (Molecular Devices) and Origin 7 (OriginLab Corporation). For TREK-1 current recordings, voltage was ramped from -120 to +45 mV within 3.5 s. Holding potential was -80 mV. For recordings of Nav1.5 current amplitudes a P/N (N=4) voltage-step protocol was used. Voltage was stepped from -80 mV to -30 ms for 50 ms. Holding potential was -80 mV. For the V_1/2_ of activation measurements, the currents were recorded with a P/N (N=4) voltage-step protocol (holding potential -80 mV) and voltage-steps of 50 ms ranging from -70 to +80 mV in 10 mV increments (see inset activation protocol). For the V_1/2_ of inactivation measurements, the currents were recorded with a P/N (N=4) voltage-step protocol (holding potential -100 mV) and voltage-steps of 50 ms ranging from -120 to -10 mV in 10 mV increments with a final step to -30 mV for 50 ms (see inset inactivation protocol). The time-course of the recovery from inactivation was recorded with a two-pulse protocol. A first step to -10 mV for 50 ms was followed by the second test pulse step to -10 mV with increasing interpulse intervals. The V_1/2_ of activation or inactivation were obtained by Boltzmann fitting.

### Statistical analysis

Statistical analysis was performed using GraphPad Prism. In case of the membrane targeting assay, data distribution was assessed using the Shapiro–Wilk test. Non-parametric comparisons were performed using the Mann–Whitney U test. In case of the BiFC assay and the two-electrode voltage-clamp recordings, data were analyzed using an unpaired two-tailed Student’s t-test. Variance equality was assessed using an F-test. All data are presented as mean ± s.e.m..

## RESULTS

### Clinical Evaluations & JET Diagnoses

All family members were diagnosed with JET at birth following 12-lead ECG in the setting of a narrow complex tachycardia. In some cases, there was a borderline tachycardia reported *in utero* (though fetal records were not available for review), though none required premature initiation of labor or premature delivery. The initial member of the family presented to our center with cJET and was managed with digoxin and quinidine for the first two years of life, followed by spontaneous resolution (Figure 1A, I-1). He was lost to follow-up until his second child with a new partner presented with cJET (Figure 1A, II-2). His first child also had cJET along with decreased LV function and was maintained on digoxin, atenolol, and flecainide (Figure 1A, II-1). Medication was stopped by the age of 5 years, showing spontaneous resolution of her arrhythmia. The second child’s rhythm was able to be converted to sinus rhythm with amiodarone and propranolol after a prolonged hospitalization in the absence of ventricular dysfunction (Figure 1A, II-2). The child tolerated the rhythm throughout and did not require additional hemodynamic support. A third child (Figure 1A, II-3), a full sibling to the second, presented with cJET and cardiovascular collapse at a referring institution. Mechanical circulatory support was required for the first 10 days and cardiac function eventually returned following rate and rhythm control with amiodarone. The cJET spontaneously resolved within the first year. All four individuals were without arrhythmia on no medication with normal heart function by echocardiogram at last follow-up. The JET diagnosis is made reliably on surface ECG with a narrow complex ECG demonstrating ventriculo-atrial dissociation (more QRS complexes than P-waves) (Figure 1B). Notably, none of the patients exhibited known or overt skeletal myopathy or symptoms suggestive of limb-girdle muscular dystrophy.

**Figure 1.**
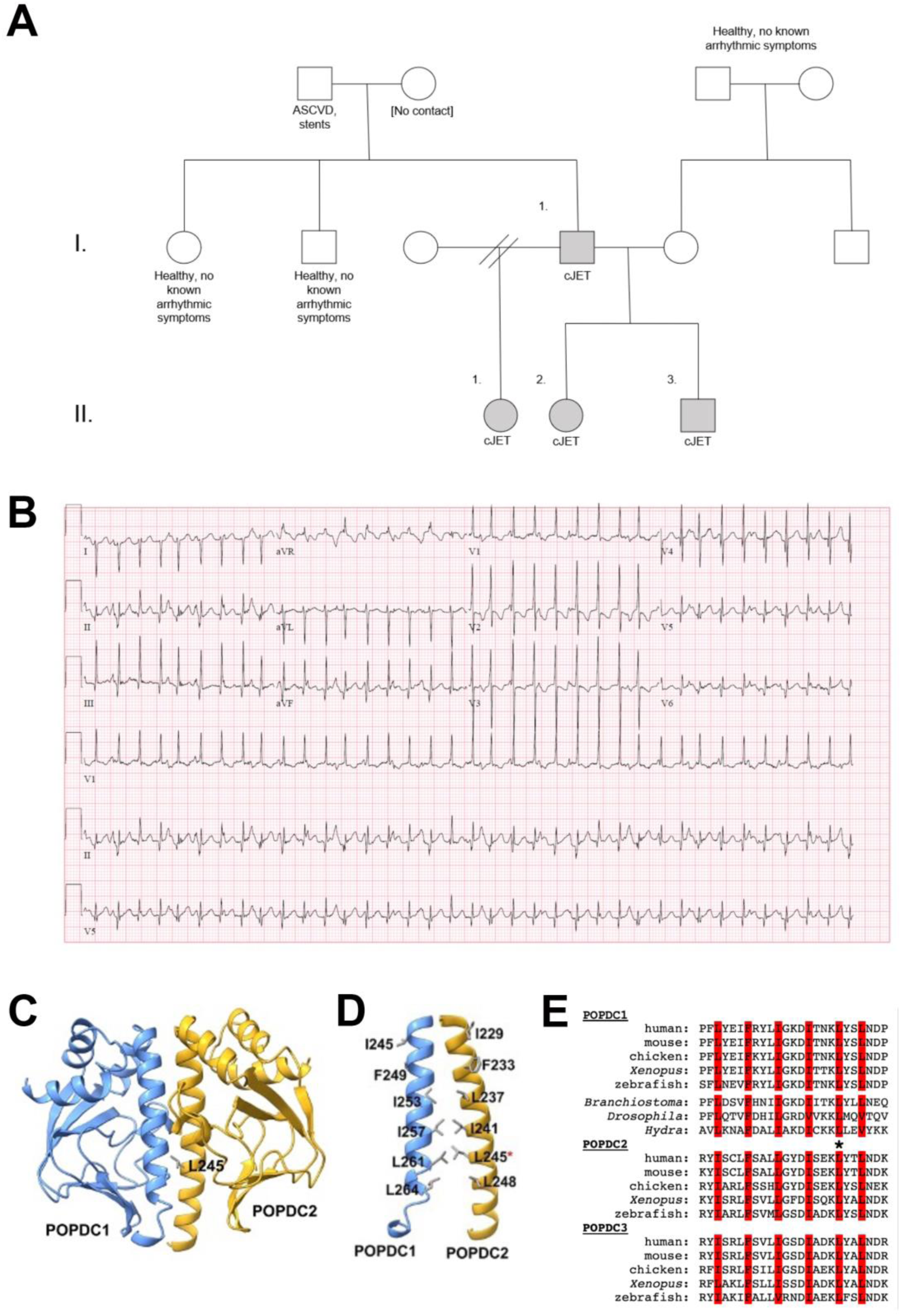
Identification of a POPDC2 p.Leu245Pro variant in a family with congenital junctional ectopic tachycardia (cJET). (**A**) Family pedigree. Details are limited to relevant information, and the grey-shaded symbols indicate the affected relatives who underwent quad exome sequencing and rare variant segregation analysis. (**B**) Electrocardiogram tracing for the affected individual II-3. Note the ventriculoatrial dissociation, which is best seen in leads II and V5. (**C**) Model showing the interaction between the Popeye domains of POPDC1 and POPDC2. The side chain of Leu245 within the αC-helix of POPDC2 is shown. (**D**) Possible interactions between the αC-helices of POPDC1 and POPDC2. The highly conserved hydrophobic residues contained within each helix, including Leu245 (red asterisk), are shown. (**E**) Protein sequence alignment of the αC-helix of vertebrate and invertebrate POPDC proteins. Conserved hydrophobic residues, which are thought to mediate heterodimer formation, are labelled in red. Leu245 in human POPDC2 is labelled with an asterisk.

### Exome Sequencing Results

All four cJET patients were subject to whole-exome sequencing. A total of 643 variants passed quality filtering thresholds, and this list was reduced to 450 variants after filtering based on genes with established Mendelian inheritance patterns. Only five genes had Exomiser scores ≥0.80 and were associated with autosomal dominant inheritance but variably seen in affected family members. Notably, there were no variants occurring in the *TNNI3K* gene, and we confirmed with the clinical laboratory that it had sufficient coverage and depth (100% at ≥20x depth). We prioritized variants that were present in all relatives with cJET. Supplemental Table S1 summarizes the list of prioritized candidate gene variants. Only one variant with Exomiser score >0.80 was seen in all affected family members—*POPDC2*: c.734T>C (p.Leu245Pro). This variant occurs in exon 3 of the *POPDC2* gene [NM_001369919.2]. The composite Exomiser score was 0.822, and additional *in silico* predictors, CADD, and REVEL scores suggested increased probability of deleteriousness or pathogenicity. At the time of analysis, the variant had very rare prevalence in gnomAD (MAF: 0.0000118). This missense variant occurs at a highly conserved position at the C-terminal part of the Popeye domain, and it was hypothesized to potentially affect the structure of the αC helix and might impair dimerization (see below for modelling and functional analysis). Exomiser prioritized this variant based on a phenotype correlation with mouse and zebrafish *Popdc2* loss-of-function mutants and morphants showing sinus bradycardia and AV-block, respectively, as well as displaying high expression levels in the cardiac conduction system (CCS) ^43,44^. As in animal models, the human heart displays high expression levels of *POPDC2* in the CCS (including the AV node) and a nonsense variant in *POPDC2*, p.Trp188* has been shown to cause AVB^45^. Moreover, POPDC proteins interact with proteins involved in cAMP signaling including adenylyl cyclase 9 ^46^ and phosphodiesterase 4 (PDE4), which are involved in heart rate modulation ^47^. POPDC2 also interacts with ion channels (e.g., TREK1 and Nav1.5) and affects their plasma membrane trafficking ^21,22^. There are also other variants co-segregated with cJET in all family members, but these were deprioritized based on population prevalence, low/no expression in the heart/CCS, Exomiser scores <0.75, and benign/tolerated *in silico* predictions. This guided us in assessing the functional effects of the *POPDC2* p.Leu245Pro variant.

### The *POPDC2* p.Leu245Pro variant affects an ultra-conserved residue located in the αC helix of the Popeye domain

It has been proposed that the concurrent loss of POPDC1 and POPDC2 expression in the plasma membrane of skeletal muscle of patients and animal models expressing POPDC1 variants may be due to a disruption of POPDC1-POPDC2 interactions ^23^. Variants that alter plasma membrane expression of POPDC proteins in biopsy material of patients have been shown to impair plasma membrane localization and to disrupt POPDC1-POPDC2 complex formation when expressed in HEK293 cells ^23^. The αC–helix, has been suggested to mediate the interaction of POPDC1 and POPDC2 ^23^. Leu245 of POPDC2, which is substituted by proline in the variant found in family members suffering from cJET, is one of six evolutionarily conserved hydrophobic residues in the αC-helix of POPDC2 (Figure 1C,D) ^23^. Leu245 is super-conserved, and the same amino acid is present at this position in both vertebrate and invertebrate POPDC proteins, suggesting an essential role for protein function (Figure 1E). For a subset of these hydrophobic residues within the αC-helix of POPDC2, site-directed mutagenesis to aspartic acid results in a strong impairment of POPDC1-POPDC2 complex formation and loss of POPDC1 and POPDC2 plasma membrane targeting in HEK293 cells ^23^. Given that Leu245 is found in the αC-helix of the Popeye domain and is super-conserved, it may be implicated in POPDC1-POPDC2 interactions. However, substitution of Leu245 with aspartic acid did not alter membrane targeting of either POPDC1 or POPDC2 in HEK293 cells ^23^.

### Molecular dynamics simulations show distortion of the POPDC1-POPDC2 interface in the POPDC2 p.Leu245Pro variant

After energy minimization, the POPDC2 p.Leu245Pro variant shows the classical effect of proline as a helix breaker ^48^, distorting the preceding helical turn (Figure 2A), which is not seen either in the case of wild-type POPDC2 nor in the case of POPDC2 p.Leu245Asp^23^. During the Molecular Dynamics (MD) simulation of the POPDC1/POPDC2 heterodimer complex, this distortion remains present, leading to the almost complete loss of the interaction of Leu261 of POPDC1 with Pro245 in POPDC2 (Figure 2B,B’,C,C’) while standard control parameters confirm that the MD simulations proceeded as would be expected (Fig. S1-S4). This contact has been recently identified amongst the highly conserved residues in the αC-helix ^23^. In addition, the kink induced by Pro245 in the αC-helix leads to a loss of the last hydrophobic contact – formed by Thr247 and Leu248 in the αC-helix with Leu174 and His175 - between the αC-helix and the rest of POPDC2. As a result, the interface between POPDC1 and POPDC2 is significantly weakened and undergoes large amplitude fluctuations not seen in wildtype (Figure 2D-I; Supplementary Figure 1A,B). In contrast, the POPDC2 p.Leu245Asp mutant maintains as much hydrophobic contact as possible between POPDC1 Leu261 and POPDC2 Asp245 where the CH_2_ group of the latter contacts the sidechain of POPDC1 Leu261 for most of the MD trajectory. Consequently, the wild-type geometry is well maintained in the POPDC2 p.Leu245Asp mutant throughout the MD simulation, in good agreement with previous experimental work ^23^.

**Figure 2.**
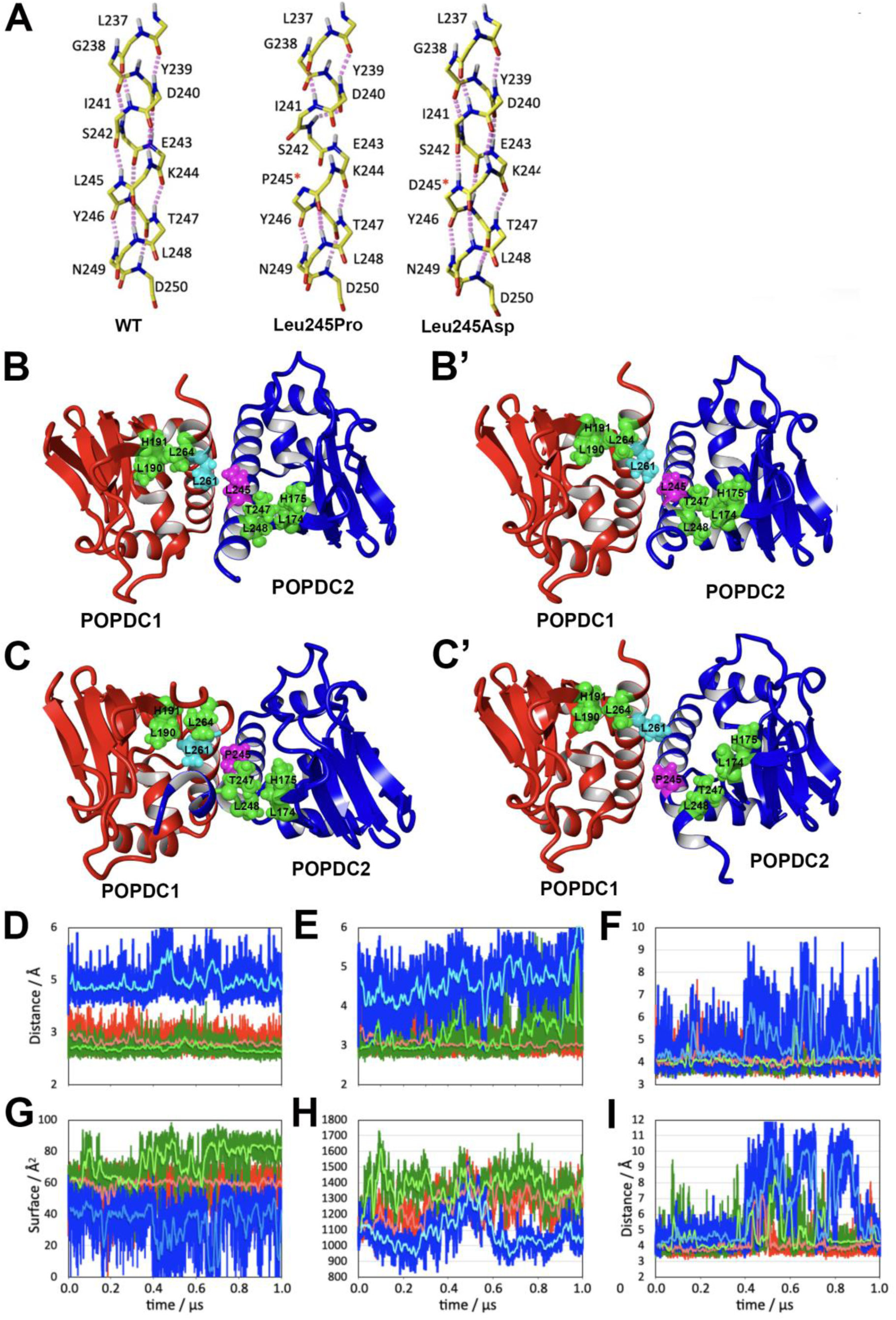
Distortion of the αC-helix in POPDC2 p.Leu245Pro leads to weakening of the dimer interface. (**A**) Detailed view of the POPDC2 αC-helix after energy minimization in wild-type POPDC2 left), POPDC2 p.Leu245Pro (middle) and POPDC2 p.Leu245Asp (right). Backbone atoms are shown in yellow (carbon), blue (nitrogen), red (oxygen) and hydrogen (white). H-bonds are indicated by magenta dotted lines. (**B,C**) Structure of the heterodimer of (**B,B’**) POPDC1-POPDC2 and (**C**,**C’**) POPDC1–POPDC2 p.Leu245Pro at the start (**B**,**C**) and after (**B’**,**C’**) 1μs of MD simulation. In addition to the main interface, contact Leu261 (POPDC1; cyan) and Leu/Pro245 (POPDC2; magenta), two clusters (green) are highlighted, marking the last hydrophobic contact of the C-terminus of the αC-helix with the rest of the protein. In POPDC1, this is Leu264 on the αC-helix, which contacts Leu190 and H191 on the proximal β-strand. For POPDC2, these are Leu248 and Thr247 on the αC-helix, which contacts Leu174 and His175 in the proximal β-strand (shown in some greater detail in Fig. S10). (**D**-**I**) Measurements are shown for a 1μs MD simulation trajectory of wild type (green), Leu245Pro mutant (blue) and Leu245Asp mutant (red). Dark colors correspond to individual time points (100ps), light colors are 100 point averages. (**D**) Distance of O241-N245 (corresponding to a helical i,i+4 H-bond). (**E**) Distance of O242-N246. (**F**) Shortest distance between sidechain atoms of Leu261 in POPDC1 and Leu/Pro/Asp245 in POPDC2. (**G**) Solvent accessible surface covered by the contact of Leu261 in POPDC1 and Leu/Pro/Asp245 in POPDC2. (**H**) Solvent accessible surface covered in the interface between POPDC1 and POPDC2. (**I**) Shortest distance between sidechain atoms of Thr247 and His175 in Leu/Pro/Asp245in POPDC2. Detailed descriptions for all of the measurements in D-I are provided in Fig. S8-S11.

### The POPDC2 p.Leu245Pro variant displays impaired interaction with POPDC1 and interferes with plasma membrane localization in HEK293 cells

To determine if the POPDC2 p.Leu245Pro variant caused a disruption of POPDC1-POPDC2 complex formation and plasma membrane localization, cDNA constructs of POPDC1 and POPDC2 p.Leu245Pro, each possessing ECFP and EYFP C-terminal tags, respectively, were co-expressed in HEK293 cells. Co-expression of wild-type POPDC1-ECFP and wild-type POPDC2-EYFP served as a positive control, where both proteins are expected to be enriched at the plasma membrane. Plasma membrane localization of each construct was quantified as the ratio of fluorescence intensity at the plasma membrane relative to the cytoplasm. As previously reported, the median plasma membrane-to-cytoplasm ratio was 4.461 (IQR: 2.937–6.039) for wild-type POPDC1 and 5.666 (IQR: 3.095–7.176) for wild-type POPDC2 when co-expressed (n=46 cells; Figure 3A,B) ^23^. When the POPDC2 p.Leu245Pro variant was co-expressed with wild-type POPDC1, there was a significant reduction in plasma membrane localization of both proteins. The POPDC1 ratio decreased to 1.308 (IQR: 0.9285–1.967; n = 34; Mann–Whitney U = 157, p < 0.0001) and the POPDC2 p.Leu245Pro ratio was 1.195 (IQR: 0.9285–1.967, U=106, p<0.0001) (n=34; Figure 3A,B). Analysis of the absolute plasma membrane fluorescence of POPDC1-ECFP in the plasma membrane of cells (normalized to the wild-type pair) showed a drop from 1.000 (IQR: 0.6621–1.431) to 0.7085 (IQR: 0.4652-0.9403, U=531, p=0.0142) when co-expressed with POPDC2 p.Leu245Pro (Supplementary Figure 12). However, there was no significant change in the concentration between wild-type and POPDC2 p.Leu245Pro at the plasma membrane (U=632, p=0.1464). However, cytoplasmic accumulation of POPDC1 and POPDC2 p.Leu245Pro was observed when co-expressed. Normalized cytoplasmic levels of POPDC1 increased 2.194-fold (IQR: 1.517–3.210, U=311, p<0.0001), and POPDC2 p.Leu245Pro levels 3.018-fold (IQR: 1.679–5.919, U=249, p<0.0001) compared to wild-type POPDC2 (Supplementary Figure 2). This shows that POPDC2 p.Leu245Pro led to the accumulation of both POPDC isoforms within the cytoplasm of the cells, which drove the loss of plasma membrane-specific expression, but had a lesser effect on the absolute levels at the plasma membrane.

**Figure 3.**
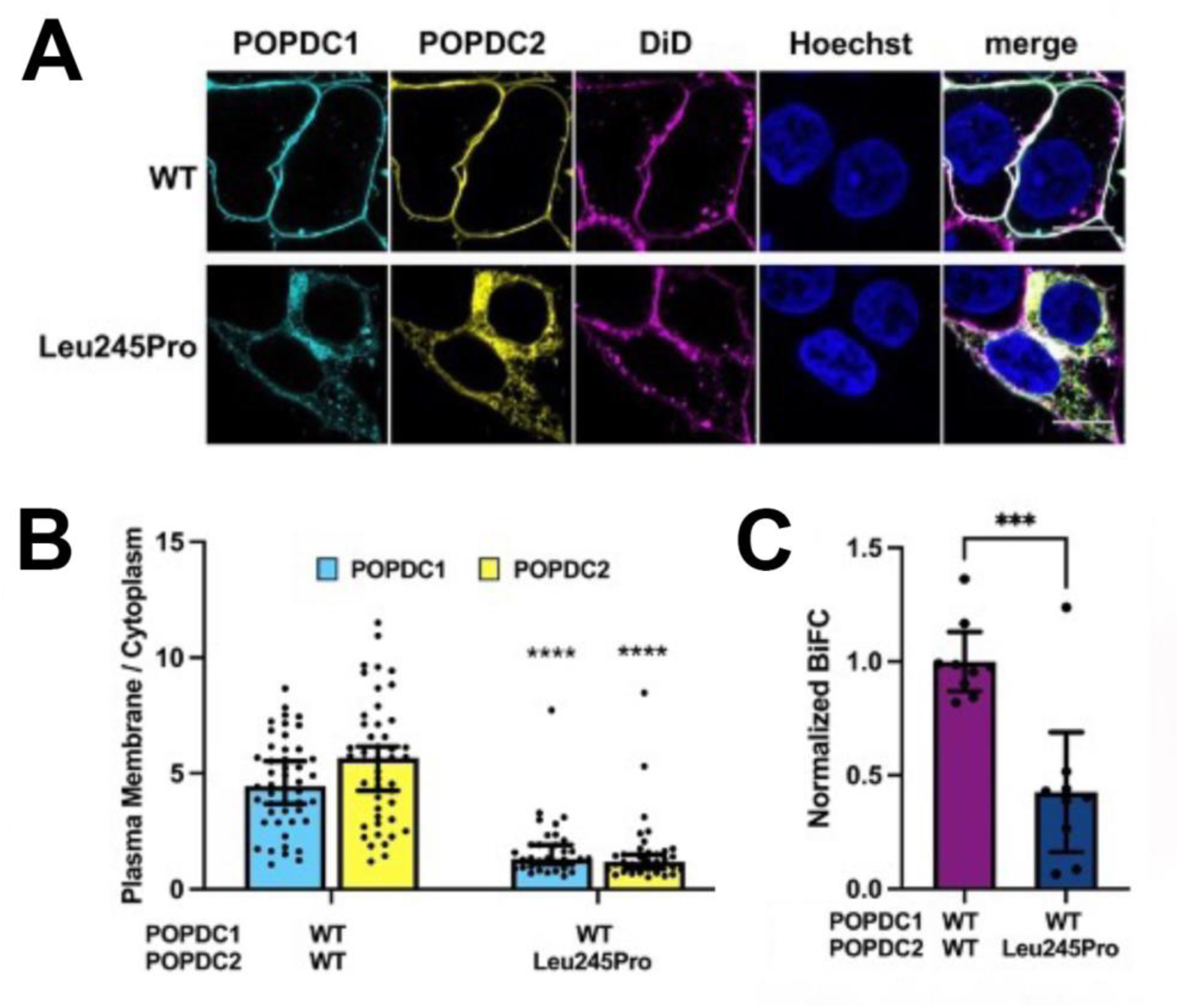
The POPDC2 p.Leu245Pro variant displays impaired membrane localization. (**A**) Representative images of HEK293 cells expressing POPDC1-ECFP and wild-type (WT) POPDC2-EYFP or POPDC2 p.Leu245Pro-EYFP. (**B**) The median plasma membrane to cytoplasm ratio of the expression of POPDC1 and POPDC2 after co-transfection of POPDC1-ECFP and POPDC2-EYFP (4.461 (IQR: 2.937, 6.039) and 5.666 (IQR: 3.095, 7.176), respectively, n=46) or POPDC1-ECFP and POPDC2 p.Leu245Pro-EYFP (1.308 (IQR: 0.9285, 1.967) and 1.195 (IQR: 0.8074, 1.667), respectively, n=34) in HEK293 cells. The change in the plasma membrane enrichment for each isoform was analyzed using a Mann-Whitney test: U=157, p<0.0001 for POPDC1 and U=106, ****p<0.0001 for POPDC2/POPDC2 p.Leu245Pro. Error bars show median ± 95% CI. (**C**) The normalized BiFC Venus emission observed after co-expression of POPDC1-VC155 and POPDC2-VN155 (1.000; 95% CI: 0.8694, 1.130; n=9) and POPDC1-VC155 + POPDC2 p.Leu245Pro-VN155 (0.4254; 95% CI: 0.1619, 0.6889; n=9). Groups were compared using an unpaired t-test: t(16) = 4.505, ***p=0.0004. Error bars show mean ± 95% CI.

### The POPDC2 p.Leu245Pro variant alters POPDC1-POPDC2 interactions in HEK293 cells

After observing that the POPDC2 p.Leu245Pro variant induced aberrant cytoplasmic accumulation of both POPDC isoforms in HEK293 cells, we investigated whether this was due to a change in the interaction of the two POPDC isoforms, as we previously reported for other pathogenic POPDC variants cells ^23^. For this purpose, a bimolecular fluorescence complementation (BiFC) assay was utilized. Wild-type POPDC1 was tagged at the C-terminal end with a VC155 fragment, whereas wild-type POPDC2 or POPDC2 p.Leu245Pro were fused with a C-terminal VN155 fragment. Both constructs were co-expressed in HEK293 cells, and mRFP was co-transfected with the POPDC constructs to act as an internal control for transfection efficiency. Venus emission levels were normalized to levels of mRFP fluorescence. Representative images of the Venus and mRFP emission from BiFC cells are shown in Supplementary Figure 13. As expected, co-expression of POPDC1-VC155 and POPDC2-VN155 generated robust Venus fluorescence, indicating an interaction between the constructs. The Venus fluorescence observed for the wild-type pair was set to 1.000 (95% CI: 0.869–1.130; n=9; Fig. 2C). Substitution of wild-type POPDC2-VN155 with POPDC2-VN155 p.Leu245Pro resulted in a reduction in the relative Venus emission to 0.4254 (95% CI: 0.162–0.689; n=9), with unpaired two-tailed Student’s t-test demonstrating statistical significance (t(16) = 4.505, p=0.0004; F-test for equality of variance was not significant (F = 4.088, dfn = 8, dfd = 8, p = 0.0628)) (Figure 3C). However, Venus emission levels above background fluorescence were observed, suggesting that some interaction between POPDC1 and POPDC2 p.Leu245Pro was still possible.

### The POPDC2 p.Leu245Pro variant increases TREK-1 and reduces Nav1.5 current amplitudes

Since we recently described a modulation of TREK-1 and SCN5A current amplitudes by POPDC2 proteins ^21,22^, we aimed to analyze the effect of the POPDC2 mutant on those channels. The current amplitudes of the TREK-1/POPDC2 complex were slightly but significantly increased by the mutant compared to co-expression of POPDC2 wild-type (Figure 4A,B). However, Nav1.5 current amplitude was enhanced by the mutant compared to wild-type POPDC2 (Figure 4C,D). This effect was only observed under theophylline-free conditions (Figure 4D,E) and thus seems to be cAMP-dependent. However, the half-maximal voltages of activation (Figure 4F and Supplement Figure 14A) or inactivation (Figure 4G and Supplement Figure 14B) were not affected by the mutant. Also, the recovery from inactivation of Nav1.5 was not altered by the POPDC2 p.Leu245Pro variant (Figure 4H and Supplement Figure 14C).

**Figure 4.**
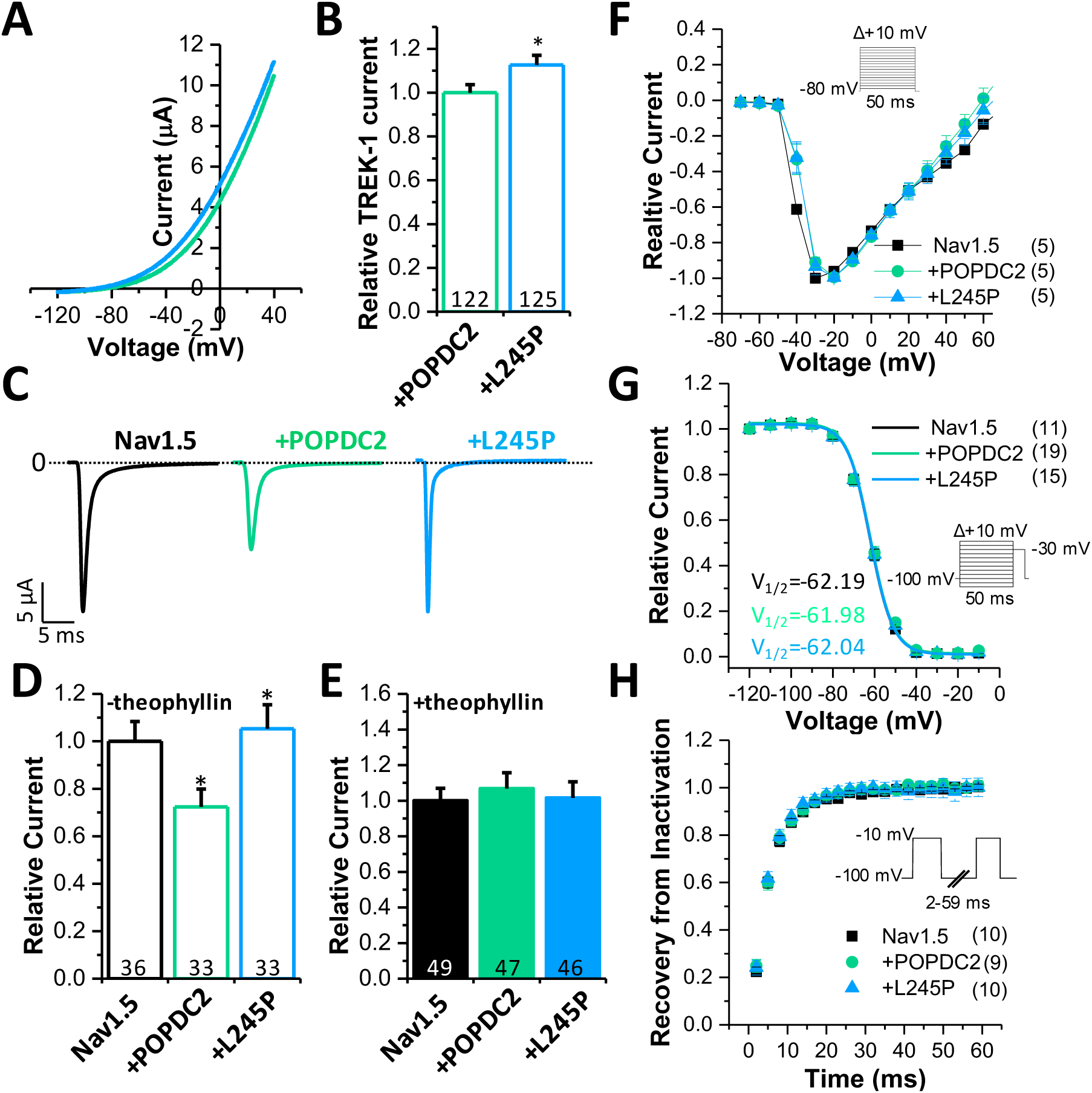
The POPDC2 p.Leu245Pro variant alters TREK-1 and Nav1.5 current amplitudes. (**A**) Representative current traces of TREK-1 (0.25 ng) co-injected with POPDC2 wild-type (1.25 ng, blue) or POPDC2 p.Leu245Pro (1.25 ng, green). (**B**) Relative current amplitudes were analyzed at +40 mV and normalized to TREK-1 + POPDC2 wild-type. The number of oocytes recorded is shown within the bars. (**C**) Representative current traces of Nav1.5 (black) Nav1.5 + POPDC2 (green), or POPDC2 p.Leu245Pro (blue), recorded by a voltage step from -80 mV to -30 mV for 50 ms. (**D**) Relative current analyzed at +30 mV and normalized to Nav1.5 without theophylline or (**E**) with theophylline in the storage solution. The number of oocytes recorded is shown within the bars. (**F**) Current voltage relationship of Nav1.5 (10 ng, black), Nav1.5 + POPDC2 (10 ng plus 10 ng, green) or Nav1.5 + POPDC2 p.Leu245Pro (10 ng plus 10 ng, blue). The number of experiments is given within the graph. Voltage protocol is shown as an inset. (**G**) Inactivation curves of Nav1.5 (black), Nav1.5 + POPDC2 (green) or Nav1.5 + POPDC2 p.L245P (blue). V_1/2_ values and numbers of experiments are shown within the graph. The voltage protocol is shown as an inset. (**H**) Recovery of inactivation of Nav1.5 (black), Nav1.5 + POPDC2 (green) or Nav1.5 + POPDC2 p.Leu245Pro (blue). The voltage protocol is shown as an inset. The number of experiments is given in brackets.

## Discussion

We report an ultra-rare clinical presentation of familial cJET in four closely related affected family members. This provided the opportunity to complete exome sequencing to identify rare genetic variants co-segregating with cJET in an autosomal dominant fashion. The genetic architecture of cJET remains elusive; however, an association of rare variants in troponin I-interacting kinase (*TNNI3K*) have been independently identified in several patients ^12,25,49,50^. In cases of postoperative JET in patients with congenital heart disease, polymorphisms of the angiotensin-converting enzyme (*ACE*) and the β1 adrenergic receptor gene (*ADRB1)* were described ^6,9^. The genetic contributions to sporadic and familial cJET remain incompletely understood due to the ultra-rarity of this disorder, though genetic susceptibility and morphological defects are likely contributory ^10^. Our study is the first to suggest the potential involvement of *POPDC2* in cJET. The only highly suspicious genetic variant co-segregating in all affected relatives with cJET was *POPDC2* c.734T>C p.Leu245Pro. This was an attractive candidate variant given its emerging roles in regulating membrane trafficking of critical ion channels like *KCNK2/TREK-1* and *SCN5A*, displaying high-level expression in the cardiac conduction system, involvement in the adrenergic control of cardiac pacemaking, its association with bradyarrhythmia in mice and AV-block in zebrafish morphants ^20,22,44,45,51^. POPDC proteins bind cAMP with high affinity and are thought to act as a novel class of cAMP effector proteins ^21,52^. We propose *POPDC2* as a candidate gene associated with cJET and tachyarrhythmia and investigated the effects of the POPDC2 p.Leu245Pro variant. The cJET condition is ultra-rare with an incidence of less than 1% of all pediatric arrhythmias and data from major electrophysiological centers over a period of 40 years identified fewer than 100 cases worldwide ^5^. The POPDC2 p.L245P variant has a prevalence of 1.18x10^-5^ in the Genome Aggregation Database (gnomAD), which suggest this allele is probably too prevalent to solely be responsible for the disease onset. We may have to assume a contribution of other small-effect alleles, which, in combination with this variant, trigger a rare tachyarrhythmia. We have not been able to identify any additional candidate gene, which may act in concert with *POPDC2* to induce the cJET condition in this family. None of the variants identified received a sufficiently high Exomiser score to be considered. Unfortunately, despite our attempts, we so far were unable to identify *POPDC2* variants in other cJET cases.

Previous studies reported a loss of plasma membrane trafficking for several *POPDC1* variants ^23,43,53–57^. These studies utilized skeletal muscle biopsy material, as patients carrying these variants develop limb-girdle muscular dystrophy R25 (LGMDR25), and taking biopsies is a routine diagnostic procedure in the case of these diseases. In contrast to *POPDC1*, no skeletal muscle abnormalities have been described in patients carrying a POPDC2 p.Trp188* variant ^45^, and this seems also to be the case for the patients carrying the POPDC2 p.Leu245Pro variant. This is a surprising and unexplained finding as *POPDC2* is also expressed in skeletal muscle, albeit at lower levels than in the heart^58^. Some weakly elevated creatinine kinase levels indicative of a subclinical myopathic process were described in patients carrying the POPDC2 p.Trp188* variant ^45^, Possibly, the lack of a severe skeletal muscle pathology is related to the fact that *POPDC3* is expressed in skeletal muscle but very little in the heart. It may functionally substitute *POPDC2* in skeletal but not in cardiac muscle, which may explain why, in patients with *POPDC2* variants, only heart disease is present.

Due to the lack of a skeletal muscle phenotype in our patients, muscle biopsies were not available to study the membrane localization of POPDC1 and POPDC2 in native muscle tissue. To nonetheless demonstrate the pathogenicity of the POPDC2 p.Leu245Pro variant, we utilized a recently published co-expression assay in HEK293 cells ^23^. To this end, POPDC1 and POPDC2, when expressed individually, are mainly localized in the cytoplasm — however, upon co-expression both proteins are trafficked to the plasma membrane. Our previous investigation found that when pathogenic variants of POPDC1 or POPDC2 were co-expressed with the partnering POPDC protein, the extent of plasma membrane localization in HEK293 cells correlated well with the pattern observed in muscle biopsies cells ^23^. While large amounts of both POPDC1 and POPDC2 p.Leu245Pro accumulated in the cytoplasm of the co-transfected HEK293 cells, the impact on plasma membrane concentrations was isoform-specific. Around a 30% drop in median POPDC1 fluorescence in the plasma membrane was observed, but POPDC2 p.Leu245Pro was present at levels seen for wild-type POPDC2. In our previous investigation of patients and mouse samples, there was often a large drop in plasma membrane localisation of both POPDC isoforms, however some variability and mutant variant-specific differences have been observed ^23^. In previous HEK293 experiments variable effects on cytoplasmic accumulation and reduced plasma membrane expression have been observed, with the net effect of reducing specific expression at the plasma membrane ^23^. We observed an accumulation of both POPDC isoforms in the cytoplasm, suggesting that the mutant POPDC proteins probably accumulate in the ER and potentially trigger an unfolded protein response, which may be part of the pathology ^59^. Moreover, based on mutant analysis in mice, it is thought that POPDC1 and POPDC2 need to undergo heterodimer formation for proper function; neither protein alone can substitute for the lack of the other ^21,23^. Based upon the HEK293 experiment, some mutant POPDC2 protein may reach the plasma membrane, where it may exert dominant-negative effects, given that the mutant protein displayed aberrant protein function in each of our assays. The behavior of the POPDC2 p.Leu245Pro variant in HEK293 cells may not perfectly correlate with the situation in the heart, meaning that caution should be applied to these results. However, in the absence of biopsy material and the strong correlation between HEK293 experiments and patient and mouse biopsies previously reported for other POPDC variants ^23^, means this is a reasonable model system for examining the behavior of the variant in patient’s hearts. Thus, impaired protein-protein interaction as suggested by the BiFC assay and the molecular dynamic simulations as well as altered membrane trafficking or accumulation of the mutant protein and its interaction partner likely occurs in the hearts of these patients.

While in HEK293 cells, mutant POPDC protein and its partnering isoform accumulate in the cytoplasm (probably in the ER), in muscle tissue from patients expressing mutant POPDC1 variants, the protein is mainly present in immunostained vesicles ^23^ or completely absent. The dependence of membrane trafficking on the co-expression and heteromerization of POPDC1 and POPDC2 is thought to be based on the masking of a putative ER retention motif, although this has not yet been experimentally demonstrated^23^. Disruption of the αC-helix interface between POPDC1 and PODPC2 as in the case of the Leu245Pro mutant protein, may cause improper masking of the ER retention motif, preventing the mutant protein and its protein partner from being released from the ER.

Interestingly, *POPDC1* and *POPDC2* variants show different traits. All *POPDC1* variants that have been reported thus far show a recessive inheritance and only patients carrying two mutant alleles develop a disease phenotype ^23,43,53–57^. In contrast, patients carrying the *POPDC2 p.Leu245Pro* variant reported in this study, and those with a *POPDC2 p.Trp188\** variant ^45^ display a dominant trait. However, recent reports described patients carrying biallelic mutations in *POPDC2*, which were associated with severe cardiac phenotypes, including cardiac conduction disease and hypertrophic cardiomyopathy ^60,61^. Thus, in the case of *POPDC2*, both recessive and dominant traits have been observed.

Leu245 is one of six hydrophobic residues invariantly present in the αC-helix of the Popeye domain ^23^. We recently hypothesized that these residues might be involved in mediating heteromeric complex formation of POPDC proteins ^23^. This function would explain the very high level of sequence conservation of these and other residues in the αC-helix. While the true nature of this putative interaction domain remains to be further characterized, introduction of negatively charged aspartic acid residues in place of three of the conserved hydrophobic residues in either αC-helix disrupted membrane trafficking of POPDC proteins in HEK293 cells^23^. The position of Leu245 within the αC-helix of POPDC2 and its high conservation level indicate that a substitution of this residue may plausibly alter Popeye domain function, probably interfering with the interactions with POPDC1. However, experimentally substituting Leu245 with aspartic acid did not cause a significant disruption in the plasma membrane localization of either POPDC1 or the mutated POPDC2 ^23^. The different behavior of the Leu245Asp and Leu245Pro POPDC2 mutant proteins might be related to the known ability of proline residues to destabilize the helical structure of α-helices. The MD simulation did indeed confirm exactly such a distortion of the αC-helix in POPDC2, which destabilizes the interface directly by reducing the contact with Leu261 in POPDC1 and indirectly via compromising the anchoring of the αC- helix to the rest of the protein. None of this is seen in the Leu245Asp mutant which, apart from a mildly reduced interface, behaves exactly like the wild-type POPDC2, in good agreement with recent work ^23^. Thus, the POPDC2 p.Leu245Pro substitution could indeed weaken the interface between POPDC1 and POPDC2, impairing protein-protein interaction and resulting in aberrant membrane localization of POPDC1 and POPDC2. This is consistent with the results of the BiFC experiment, which suggested that the POPDC2 p.Leu245Pro variant did cause an impairment of POPDC1-POPDC2 interactions. It should be noted that POPDC1-POPDC2 interactions were not completely abolished, possibly due to the presence of additional POPDC1-POPDC2 interfaces distal to the αC-helix ^23^.

As already mentioned, a polymorphism of *ADRB1*, has been linked to cases of postoperative JET in patients with congenital heart disease ^9^. In this regard, it is interesting that POPDC proteins carry a high-affinity cAMP-binding site and interact with proteins involved in cAMP signaling, namely adenylyl cyclase 9 (AC9) and phosphodiesterase 4 (PDE4) ^21,46,47^. While the POPDC – PDE4 interaction protein does not affect the enzymatic activity of PDE4, a role in negative feedback regulation has been observed for the POPDC – AC9 protein complex ^62^. Interestingly, PDE4 binding to POPDC protein has an important regulatory function in the heart and is important for cardiac pacemaking ^47^. These data help to make a functional connection between the POPDC2 p.Leu245Pro mutant and the His-bundle tachycardia. The loss-of-function allele of *POPDC2* possibly results in enhanced AV node automaticity due to enhanced cAMP production as a result of the lack of negative feedback regulation to modulate AC9 enzyme activity. Moreover, the impact of the mutant protein on the altered membrane localization of TREK1 and Nav1.5 could also possibly affect the automaticity of the AV node.

As mentioned already, a biallelic nonsense mutation of *POPDC2* was found to be associated with ventricular tachycardia ^60^, thus, while in most cases *POPDC1* and *POPDC2* variants are associated with sinus bradycardia and AVB, tachycardia phenotypes have also been described. At birth, the sympathetic nervous system activity is high, aiding in the transition to extrauterine life, including increased blood pressure and heart rate ^63^. POPDC proteins are thought to ensure a physiological responsiveness of the heart to adrenergic signaling ^20^. Therefore, birth associated with increased sympathetic tone may represent a period of extra vulnerability for carriers of *POPDC2* mutant variants, which would explain that the cJET patients were phenotypically normal once the supraventricular tachycardia was successfully treated. Further work including modeling cJET in animal models or alternatively in cardioids might help to obtain better insight into the underlying pathophysiology.

## Data Availability

ll data produced in the present study are available upon reasonable request to the authors

## Abbreviations

AVB: atrioventricular block
BiFC: bimolecular fluorescence complementation
cJET: Congenital Junctional Ectopic Tachycardia
JET: Junctional Ectopic Tachycardia
*KCNK2*/TREK1: Potassium Channel Subfamily K Member 2/TWIK-Related K^+^ Channel
LGMD: limb-girdle muscular dystrophy
MD: molecular dynamics, POPDC - Popeye Domain-Containing Protein
SCN5A: sodium voltage-gated channel alpha subunit 5
TNNI3K: troponin I-interacting kinase
K_2P_: channel

## Acknowledgments

Expert technical assistance of Mrs. Ursula Herbort-Brand is hereby gratefully acknowledged. The Facility for Imaging by Light Microscopy (FILM), Faculty of Medicine, Imperial College London, which is in part funded by the British Heart Foundation (RE/18/4/34215), is acknowledged.

## Supplement Figures

**Supplement Figures 1.**
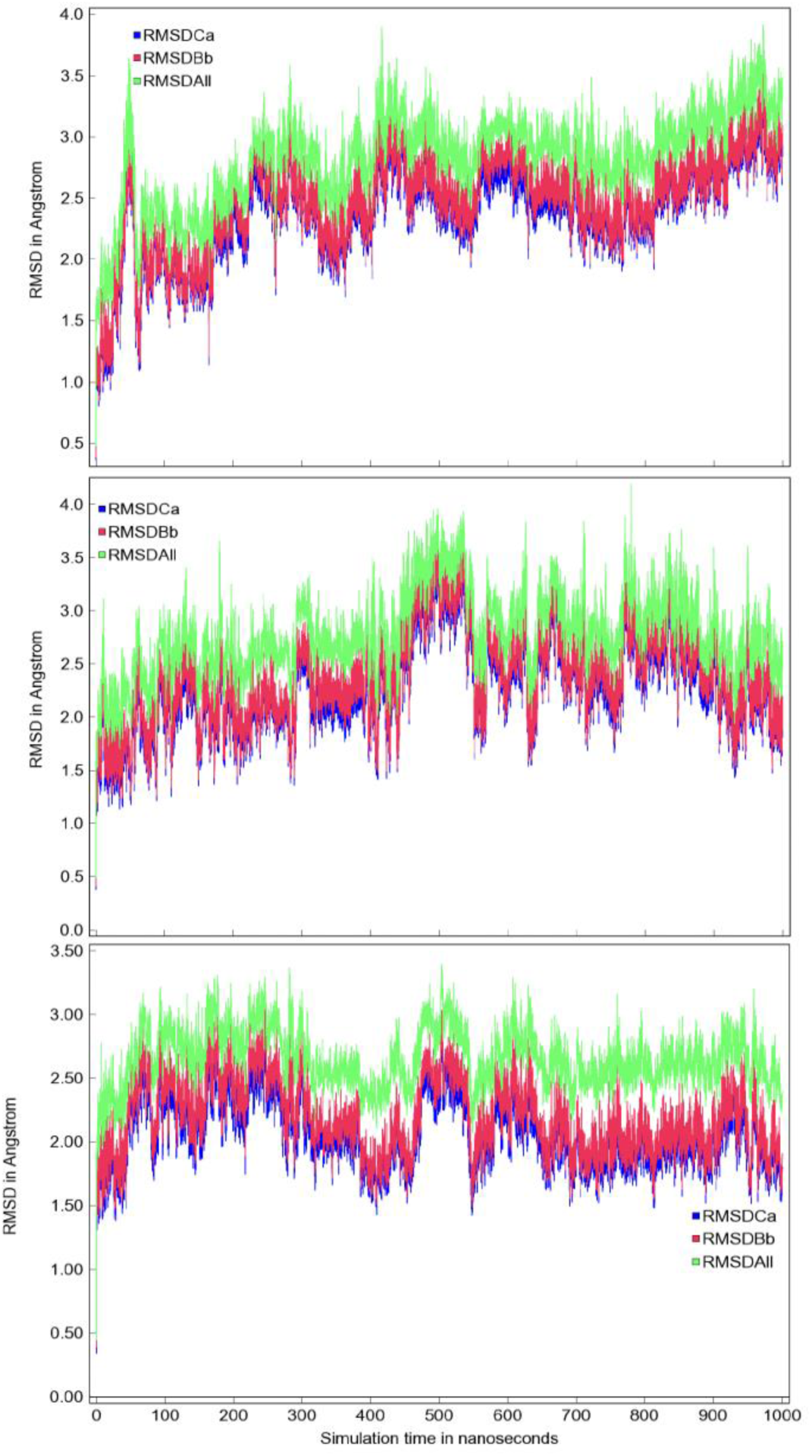
Root Mean Square Deviation (RMSD) from the starting structure for wildtype (top), L245P (middle), and L245D (bottom). There are no major overall fluctuations in any of the wild-type or mutant structures. The initial equilibration is relatively fast, taking less than ∼100 ns to reach an RMSD of ∼2-2.5 Å. Over the rest of the trajectory, all variants settle at an RMSD of 2.5-3.0 Å.

**Supplement Figure 2.**
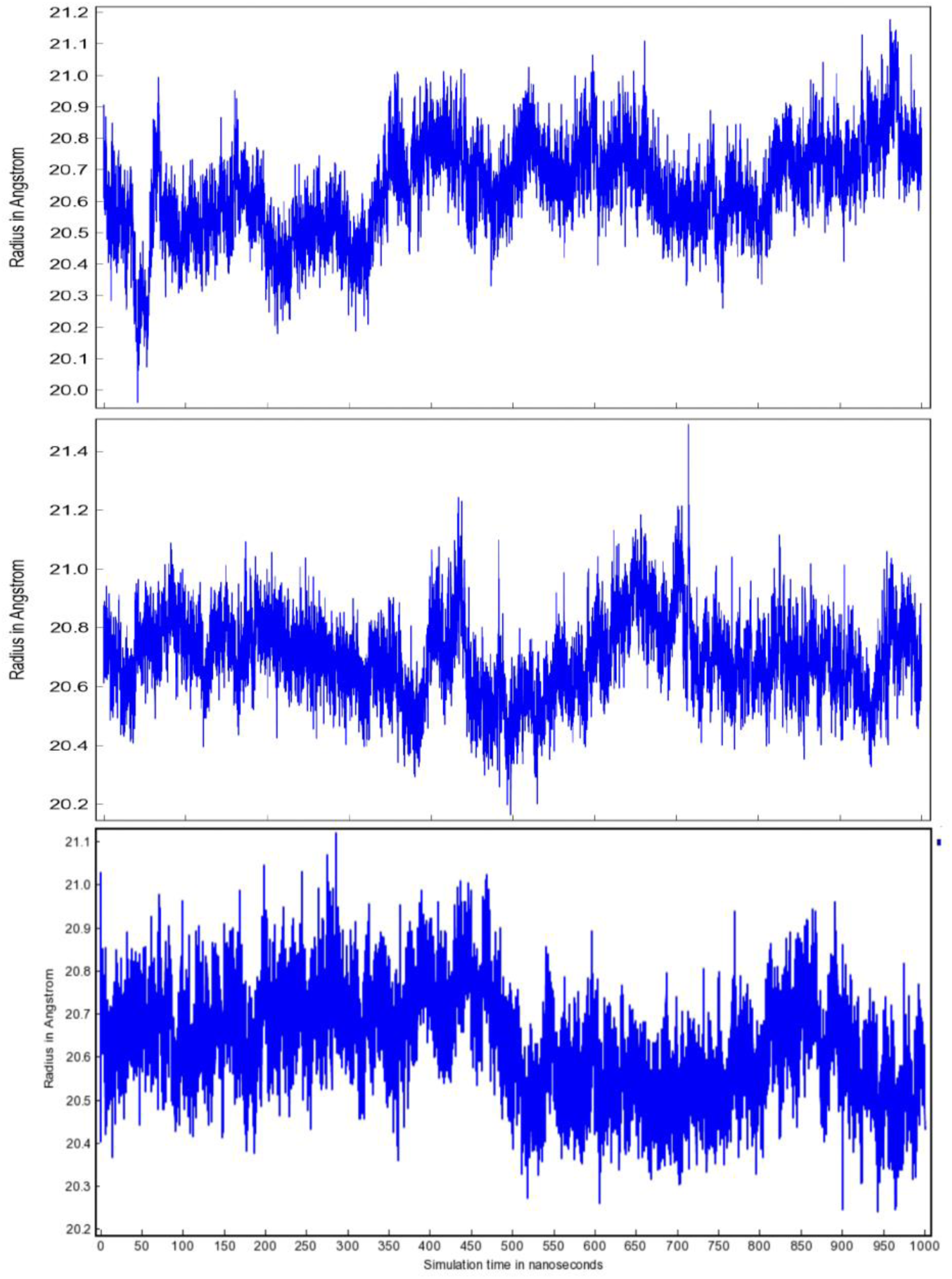
Radius of gyration for wildtype (top), L245P (middle), L245D (bottom). The apparently large fluctuations are actually very small and cover a range of 20.3-21.0 Å in both mutants and wild-type proteins. This suggests that there are no large-scale conformational changes that would affect the overall shape of the molecule.

**Supplement Figure 3.**
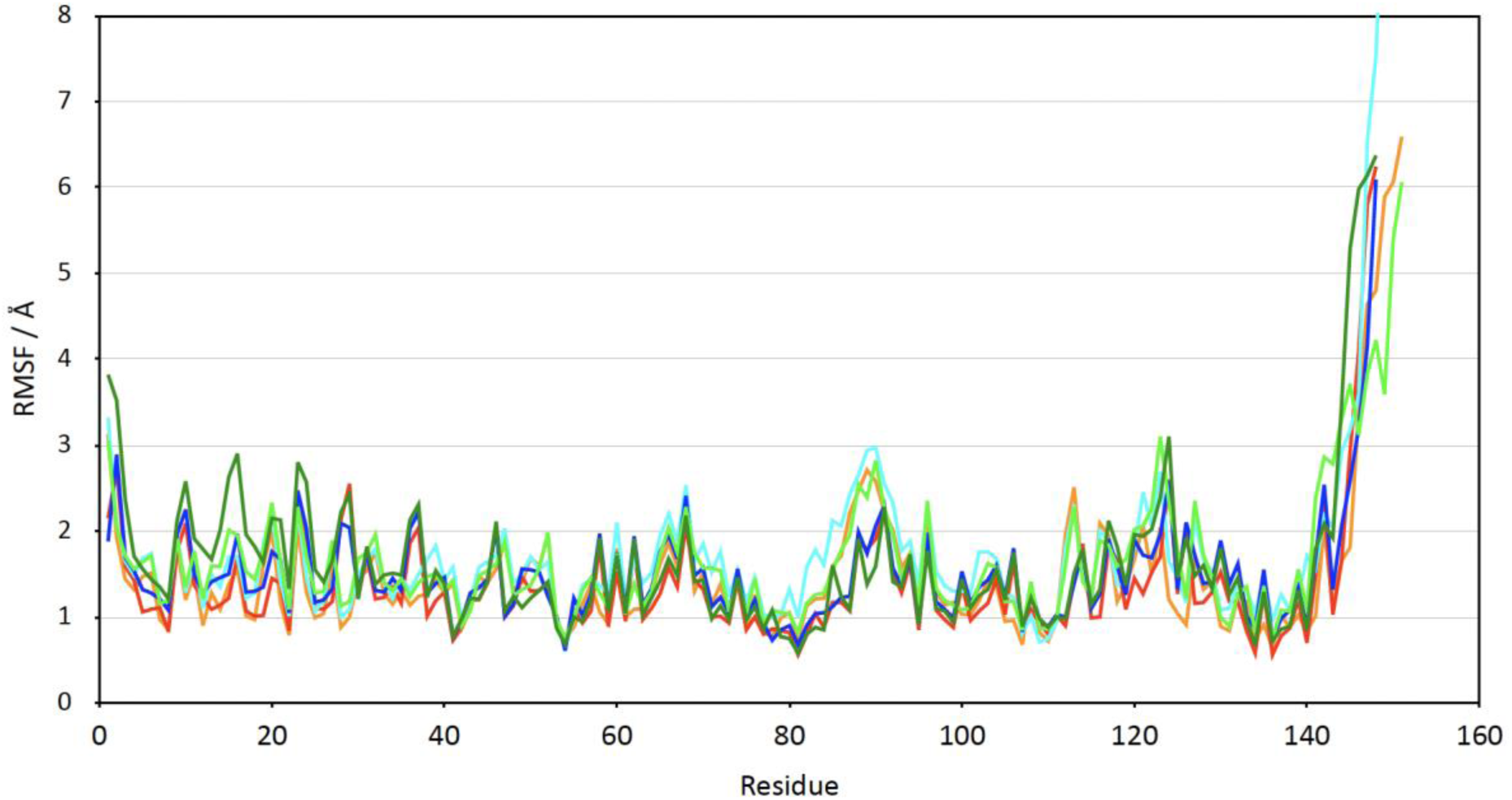
Root Mean Square Fluctuation (RMSF) values for wildtype (POPDC1: dark green, POPDC2: bright green), POPDC2 L245P (POPDC1: dark blue, POPDC2: cyan) and POPDC2 L245D (POPDC1: red, POPDC2: orange). RMSF values are quite uniform for both molecules in the heterodimer, with values around 1.5-2.0 Å. The only exception is the C-termini, where the end of the αC-helix undergoes larger amplitude fluctuations due to the shortage of close contacts. This is significantly increased in the L245P mutant due to the distortion of the helix, leading to poor packing of the rest of the protein in POPDC2. To facilitate plotting against a common X-axis, the residue numbering has been adapted. In order to reach the correct (full-length) residue numbers, a value of 120 must be added for POPDC1 and 104 for POPDC2, respectively.

**Supplement Figure 4.**
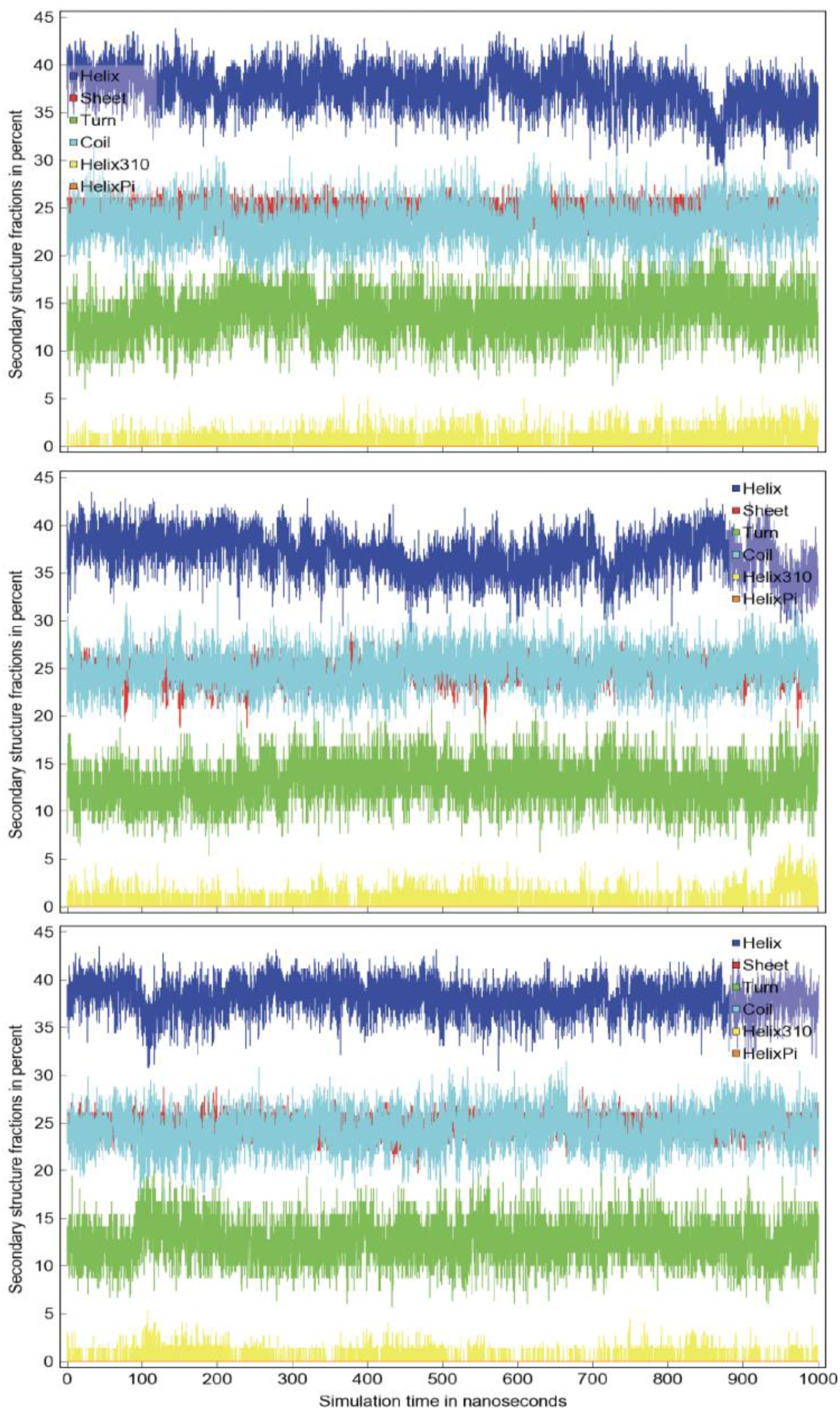
Secondary structure content for POPDC2 wildtype (top), POPDC2 L245P mutant (middle), and POPDC2 L245D (bottom). Overall secondary structure is not changing significantly during the simulation.

**Supplement Figure 5.**
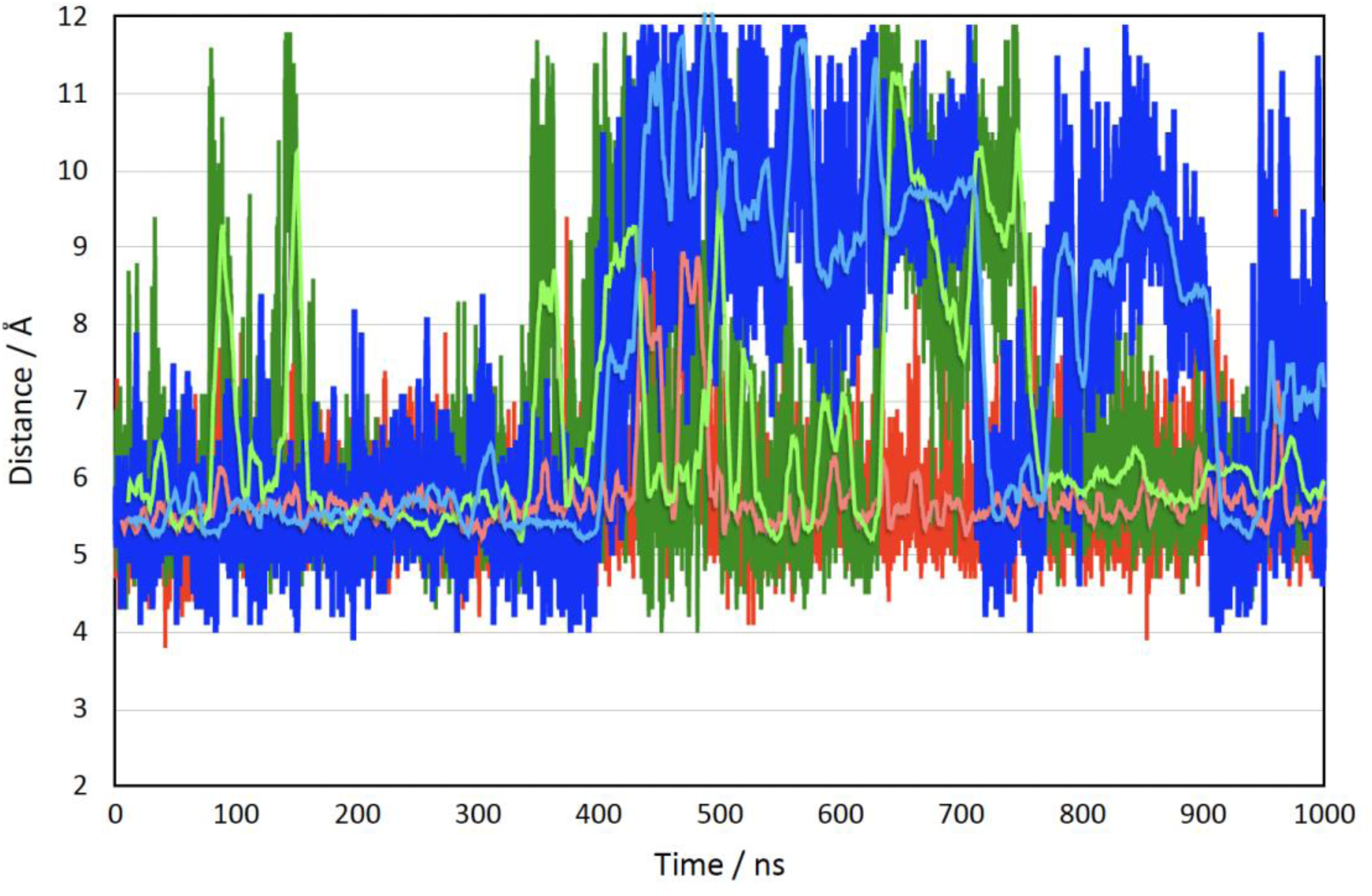
Shortest distance between the side chains of POPDC2 L248 and L174. Data for wildtype are shown in green, the POPDC2 L245P mutant in blue and POPDC2 L245D in red. The darker colors are individual time points, the lighter colors are averages of 100 time points (time points are recorded in steps of 100ps). This shows the loosening of the αC-helix contact with the rest of the protein within POPDC2 in case of the POPDC2 p.Leu245Pro mutant. The contact is undergoing quite some dynamic movement in the wildtype protein as well, but the periods of loss of contact are significantly more pronounced in the L245P mutant.

**Supplemental Figure 6.**
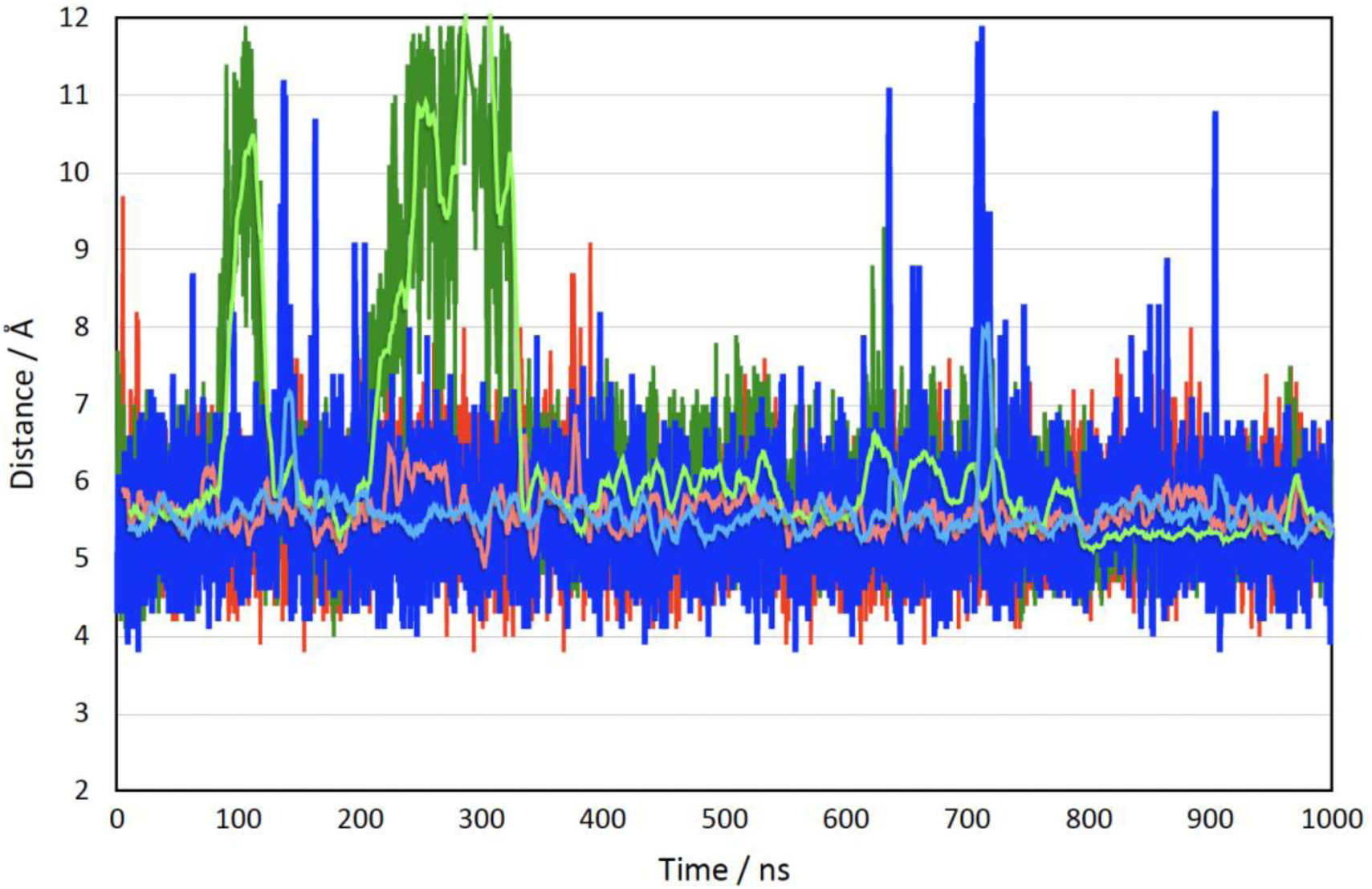
Shortest distance between the side chains of POPDC1 L264 and H190. Data generated in the presence of wild-type POPDC2 are shown in green, POPDC2 L245P mutant in blue and POPDC2 L245D mutant in red. The darker colors are individual time points, the lighter colors represent averages of 100 time points (time points are recorded in steps of 100ps). After the initial settling-in period of ∼300 ns, both wildtype and mutants remain very steady with only modest changes. This hydrophobic contact holds the αC-helix against the main body of the protein in POPDC1 and acts as a control to the corresponding contact in POPDC2, which is lost in the mutant.

**Supplemental Figure 7.**
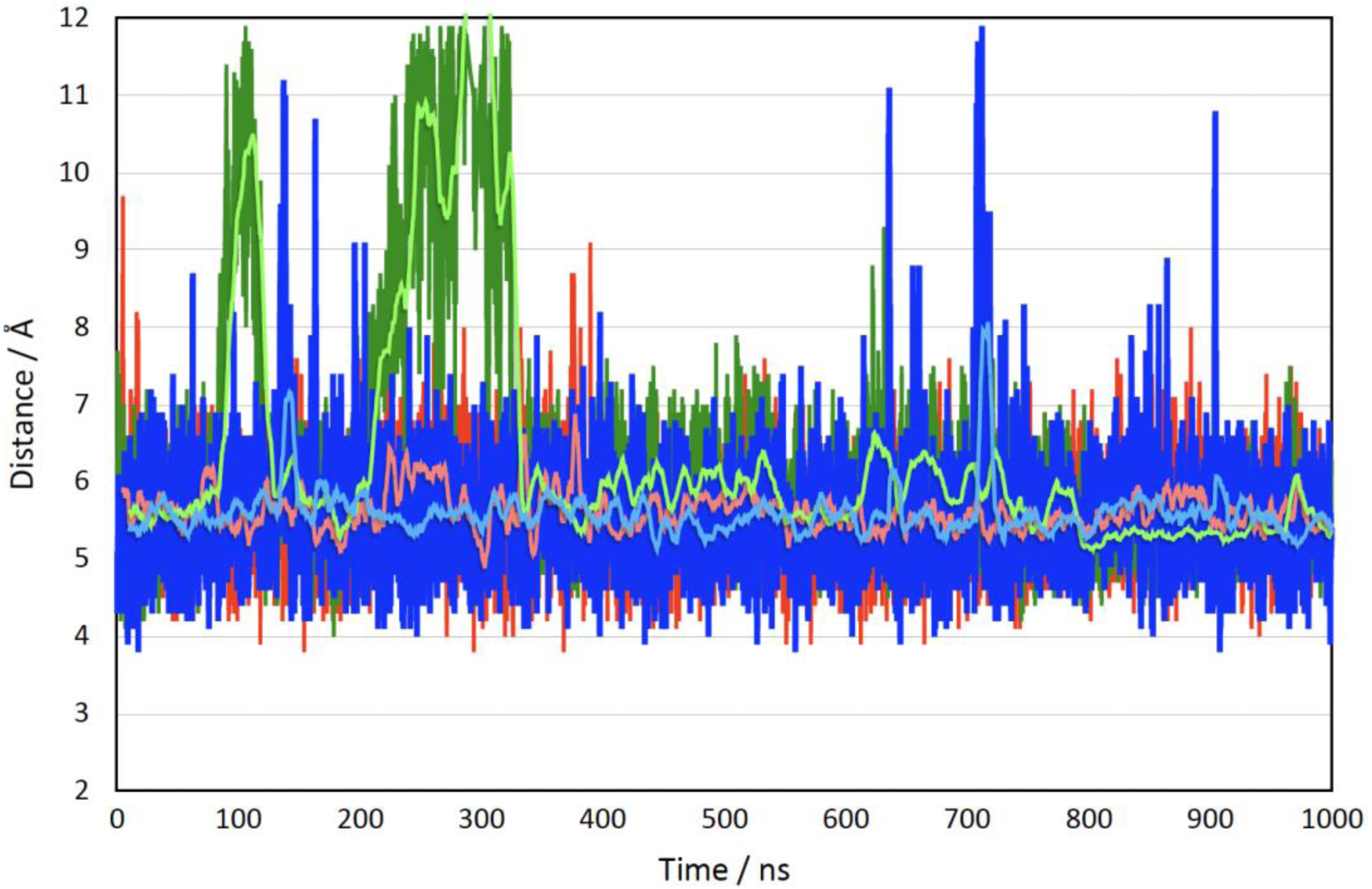
Shortest distance between the side chains of POPDC1 L264 and L191. Data for wildtype are shown in green, POPDC2 L245P mutant in blue and POPDC2 L245D in red. The darker colors are individual time points, the lighter colors represent averages of 100 time points (time points are recorded in steps of 100 ps). After the initial settling-in period of ∼300 ns, both wildtype and mutants remain very steady with only modest changes. This hydrophobic contact holds the αC-helix against the main body of the protein in POPDC1 and acts as a control to the corresponding contact in POPDC2, which is lost in the mutant.

**Supplemental Figure 8.**
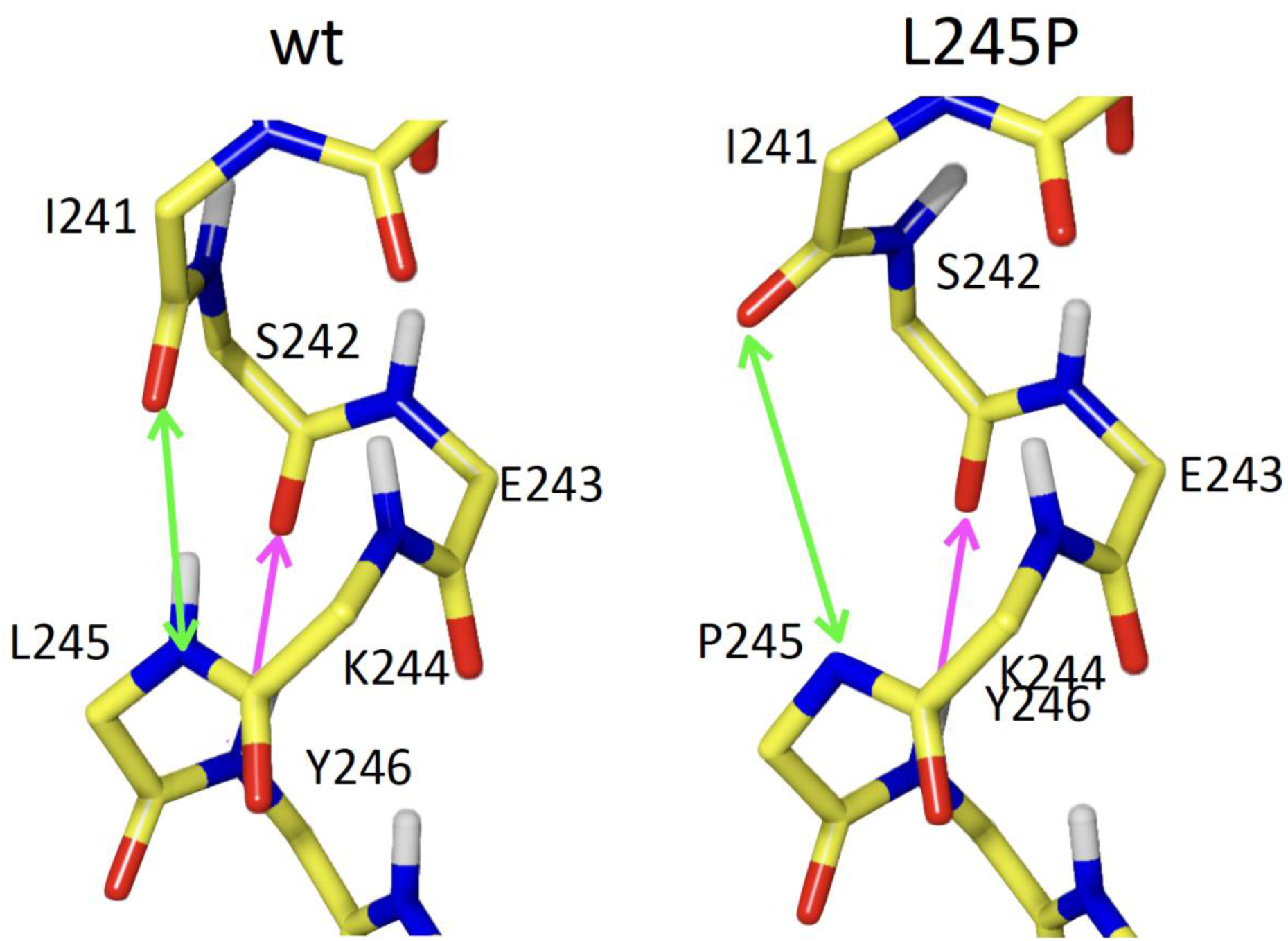
(Potential) hydrogen bond distances in the center of the αC-helix of POPDC2, around the mutated residue L245. Two distances were analyzed for the MD-simulation: distance of O241-N245 (green arrow), corresponding to the hydrogen bond between I241 and L/P/D245 (MD simulation: Figure 2E) and the distance of O242-N246 (magenta arrow), corresponding to the hydrogen bond between S242 and (MD simulation: Figure 2F). Note that the nitrogen of Y246 is partly hidden behind the oxygen of L/P245.

**Supplement Figure 9.**
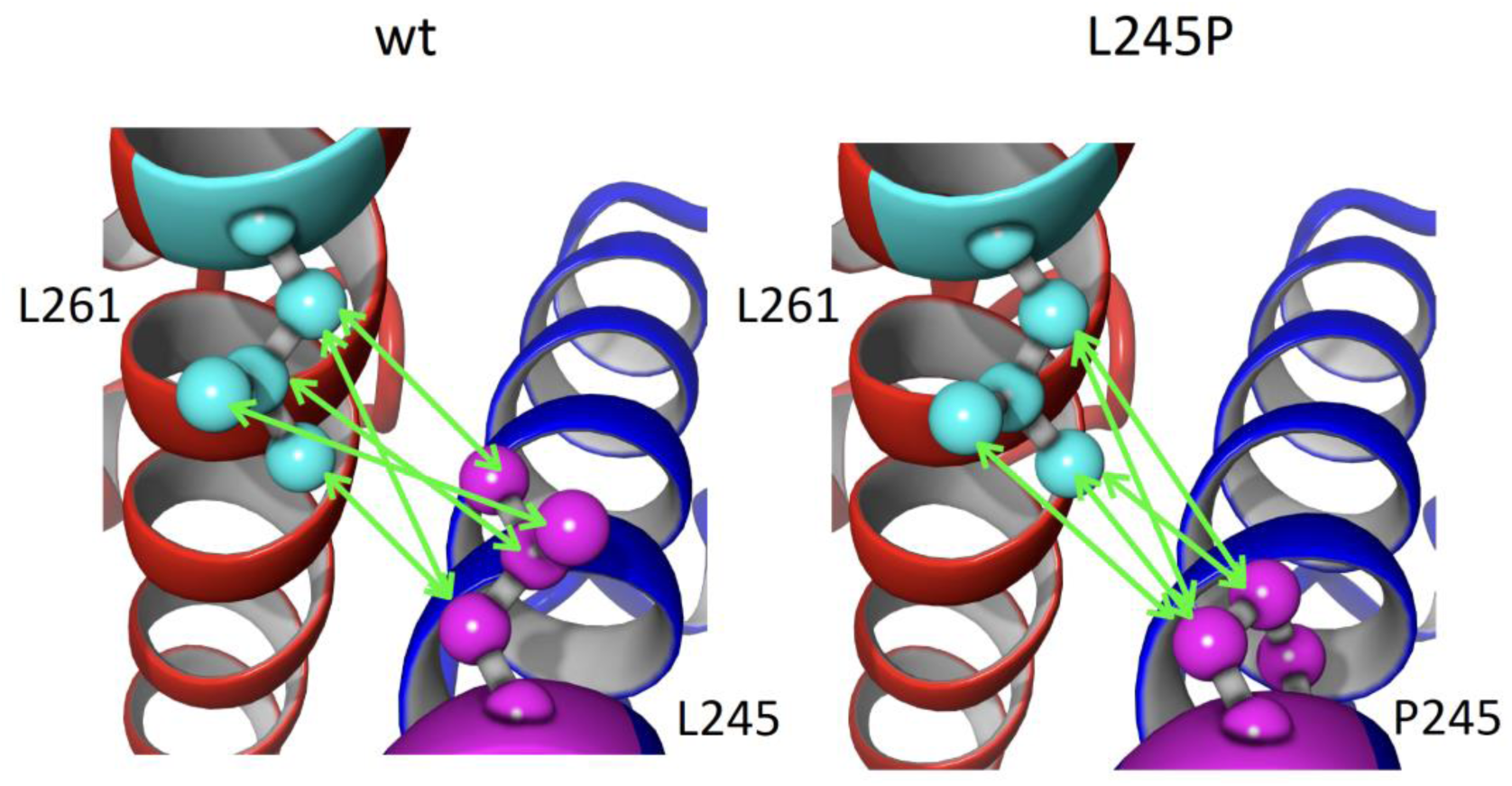
Illustration of the calculation for the distance measurement between residues POPDC1 L261 and POPDC2 L/P245, shown in Figure 2G. All carbon-carbon distances between the side chains of these two residues are measured, and then the shortest is chosen for the analysis. This ensures that, e.g. flips of the two delta methyl groups in a leucine do not accidentally give the impression of a large change in distance when in fact only one methyl group replaces the other one. Note that not all potential distances between these residue pairs are shown.

**Supplement Figure 10.**
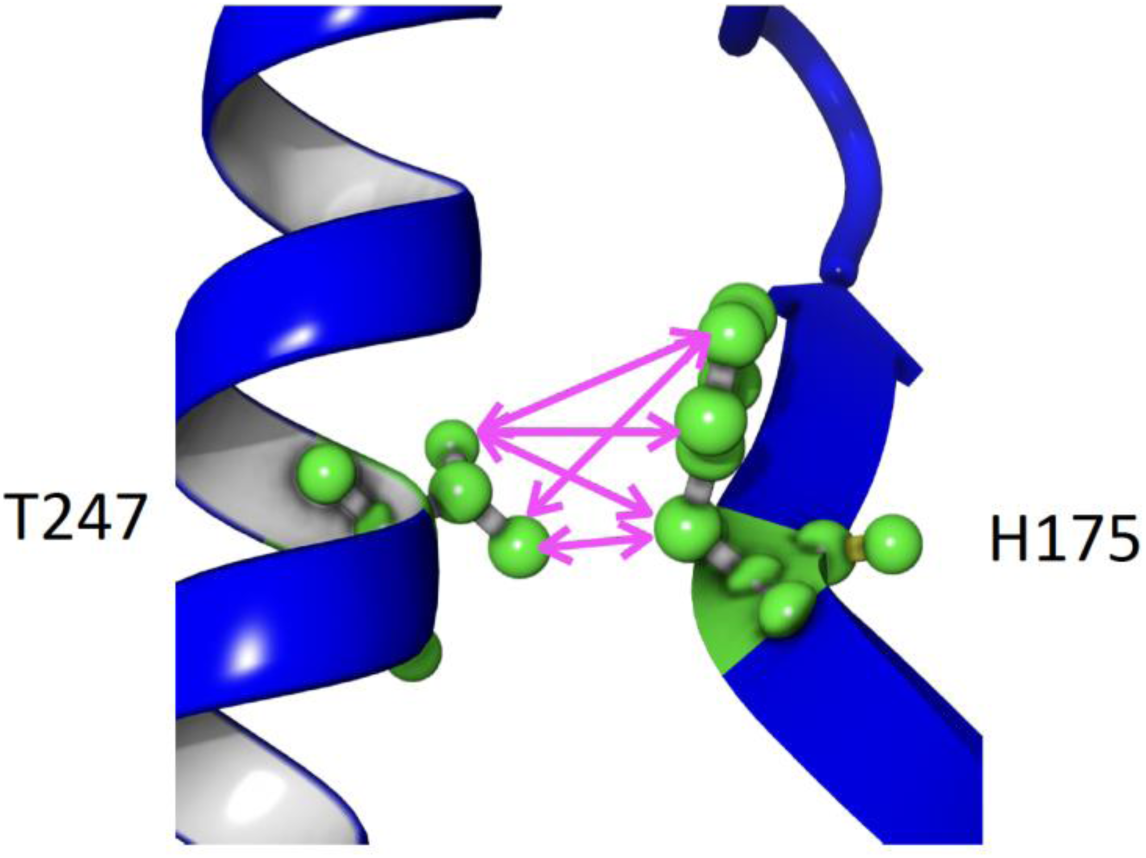
Illustration of the distance measurement calculation of residues H175 and T247 in POPDC2 (Figure 2G). This contact is part of the anchor that attaches the αC-helix to the rest of POPDC2. All carbon-carbon distances between the side chains of these two residues are measured, and then the shortest is chosen for the analysis. This ensures that, e.g. side chain rotations do not accidentally give the impression of a large change in distance. Note that not all potential distances between these residue pairs are shown.

**Supplement Figure 11.**
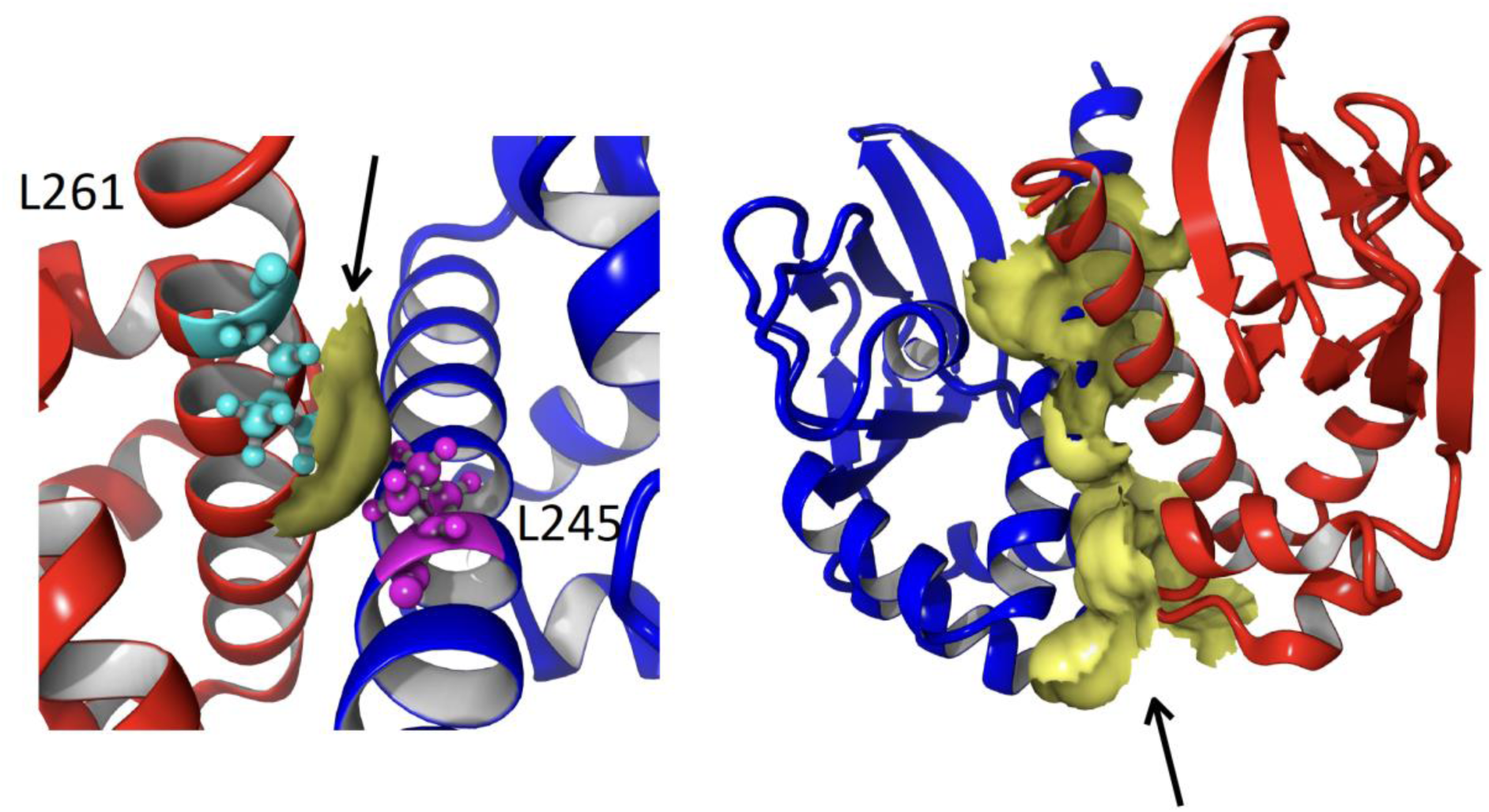
Illustration of the contact surfaces used in the analysis of the MD simulation in Figure 2. On the left, the solvent accessible surface (SAS) covered by the contact between L261 (cyan) in POPDC1 (red) and L245 (magenta) in POPDC2 (blue) is shown in yellow (corresponding to Figure 2H). On the right, the entire SAS covered by the contact between all of P1 with all of P2 is shown in yellow (corresponding to Figure 2I).

**Supplement Figure 12.**
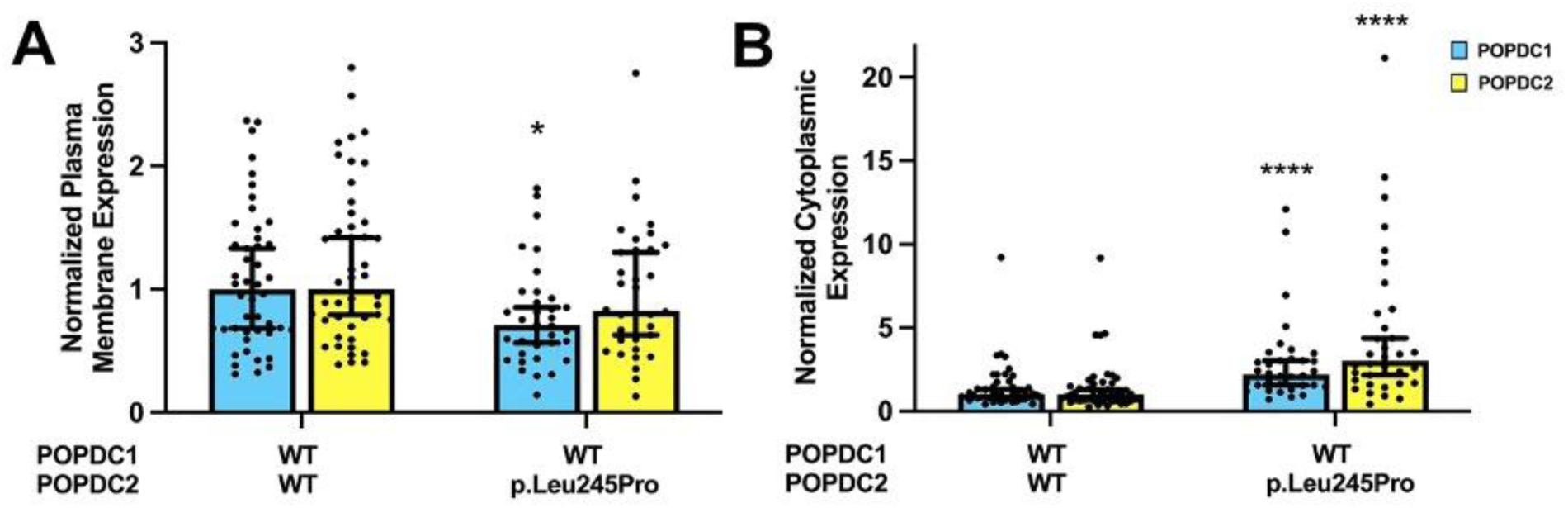
Expression of POPDC1 and POPDC2 in HEK293 cells after transfection with POPDC1 and wild-type POPDC2 or the p.Leu245Pro variant. (**A**,**B)** Normalized median expression levels of POPDC1-ECFP and POPDC2-EYFP in (**A**) plasma membrane and (**B**) cytoplasm of HEK293 cells. Bars show median ± 95% CI. The Mann-Whitney test was used to assess differences between groups. * *p* <0.05, **** *p* <0.0001

**Supplement Figure 13.**
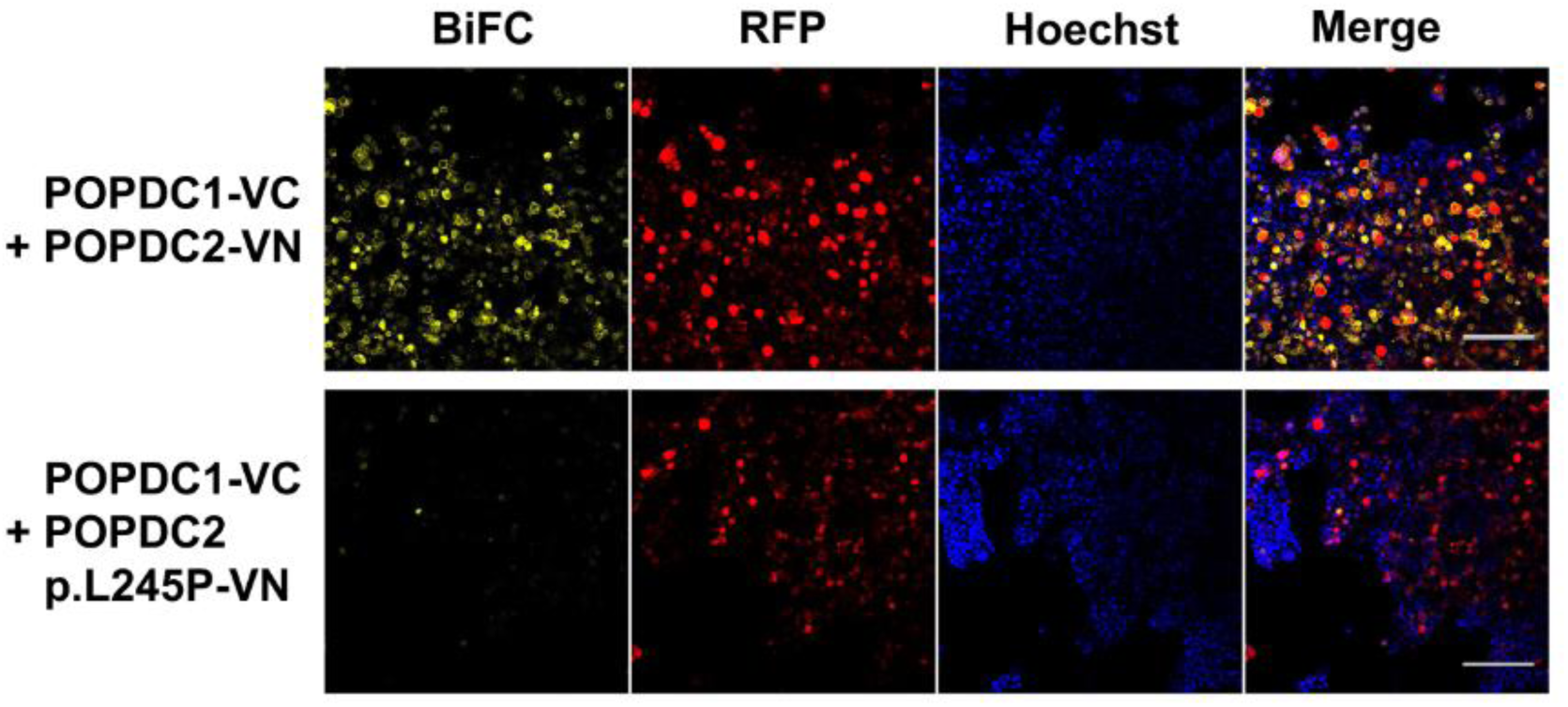
Bimolecular fluorescence complementation (BiFC) experiments. Representative confocal microscopy images of HEK293 cells transiently transfected with POPDC1-VC155 and (top panel) POPDC2-VN155 or (lower panel) POPDC2 p.Leu245Pro-VN155. Cells were also transfected with mRFP to define transfected cells and to act as internal control for Venus fluorescence. Nuclei were stained with Hoechst-33342. Scale bar: 200 μm.

**Supplement Figure 14.**
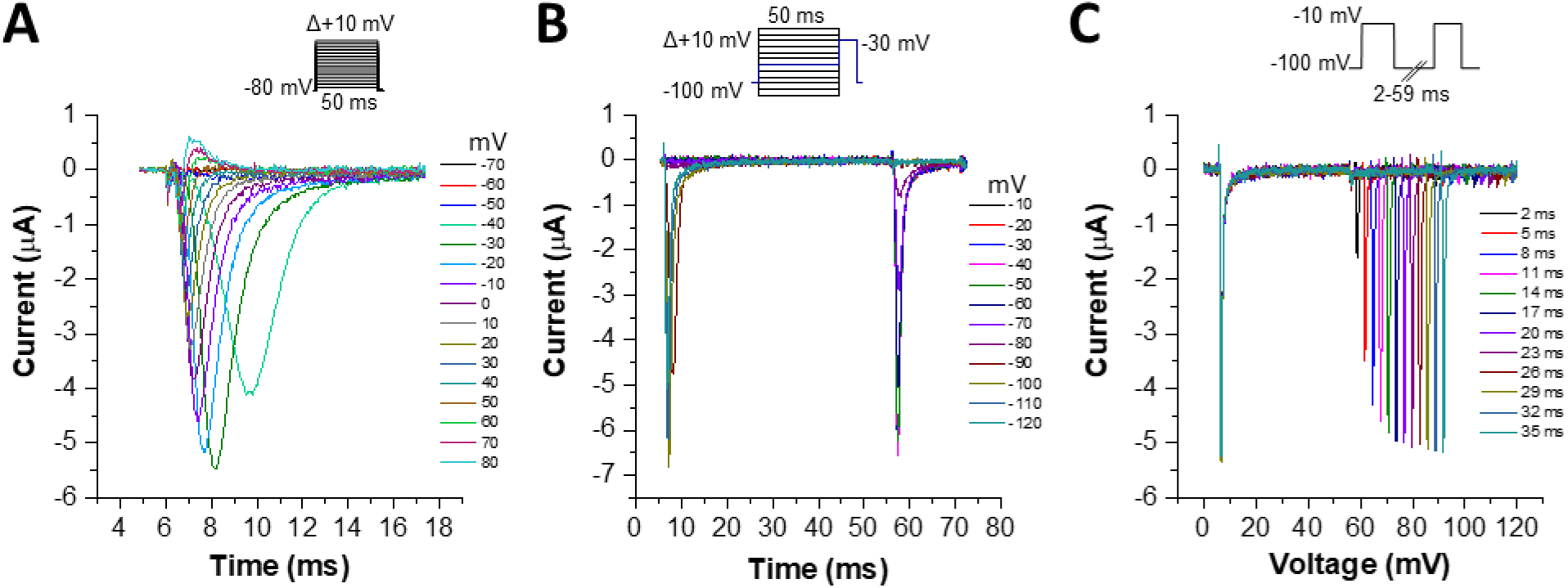
Representative original Nav1.5 current measurements. (**A**) Measurements of Nav1.5 channel activation, (**B**) inactivation, or (**C**) recovery from inactivation, applying the indicated voltage protocols.

## Legend to the Supplement Movies

### Movie 1

αC-helix of wild-type POPDC2 in the POPDC1-POPDC2 heterodimer. Only the backbone of the core part of the helix is shown to illustrate the dynamics of the typical helix hydrogen bonds between residues I,i+4. Backbone atoms are shown in yellow (carbon), blue (nitrogen), red (oxygen) and white (hydrogen). Hydrogen bonds identified by Yasara are indicated by pink dotted lines. L245 is in the centre of the helix segment.

### Movie 2

αC-helix of POPDC2 L245P in the POPDC1-POPDC2 heterodimer. Residue 245 is roughly in the centre and easily recognisable due to the missing hydrogen bond. Increased fluctuations, lack of neighbouring hydrogen bonds and bending of the helix are all evident.

### Movie 3

HHHD contact in wildtype: short segment of the MD simulation showing key contacts in the POPDC1-POPDC2 interface, as well as in the packing of the αC-helix to the remainder of the Popeye domain in POPDC1 and POPDC2. Cyan: L261 in POPDC1, pink: L245 in POPDC2, green: clusters of hydrophobic contacts holding the aC-helix to the rest of the protein. On the left is L264 in αC-helix packing to H191 and L190 in POPDC1 (red), on the right the packing of T247 and L248 in αC-helix packing to H175 and L174 in POPDC2 (blue). Apart from the occasional rearrangement, L261 in POPDC1 and L245 in POPDC2 are always very close to each other. The same applies to the two green clusters, where hydrophobic contacts between the αC-helix and the rest of the domain are very stable, both in POPDC1 and POPDC2. This video serves as a reference for the two following videos showing the same view in the POPDC2 L245P mutant protein.

### Movie 4

HH contact in the POPDC1 - POPDC2 L245P heterodimer: The focus of this video is the contact of L261 in POPDC1 with P245 in the L245P mutant of POPDC2. Colours are identical to the reference video (Movie 3). The distance between L261 in POPDC1 and P245 in POPDC2 is much larger than in the equivalent video of the wildtype heterodimer.

### Movie 5

HD contact L245P: The focus of this video is the cluster of residues T247, L248, H175 and L174 that fixes the end of the αC-helix on the rest of P2. This contact is highly mobile, much more than the equivalent in wildtype and in comparison to the symmetry-related contact L264, H191 and L190 in POPDC1 of the mutant heterodimer.

---

**Supplemental Table S1.**
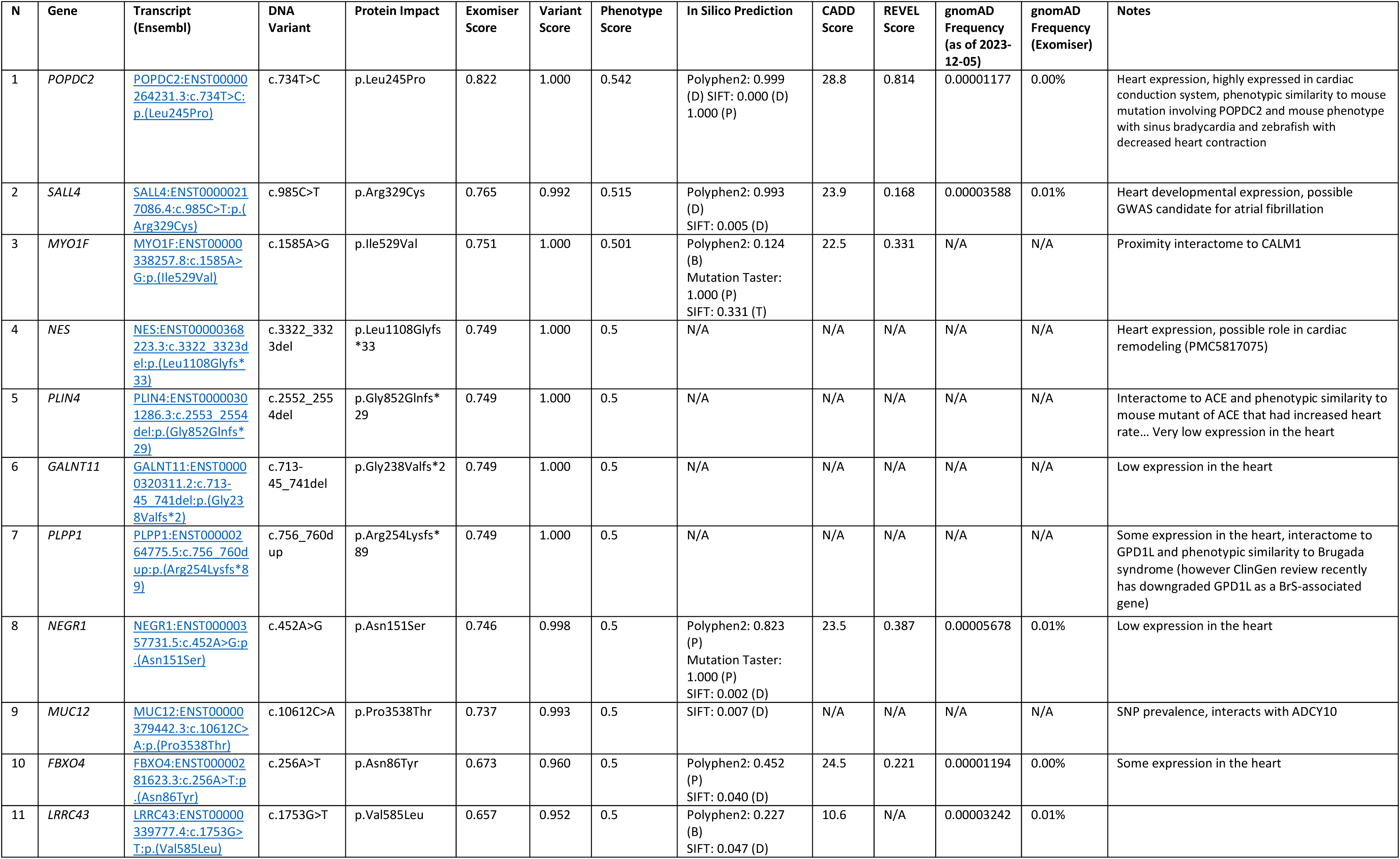
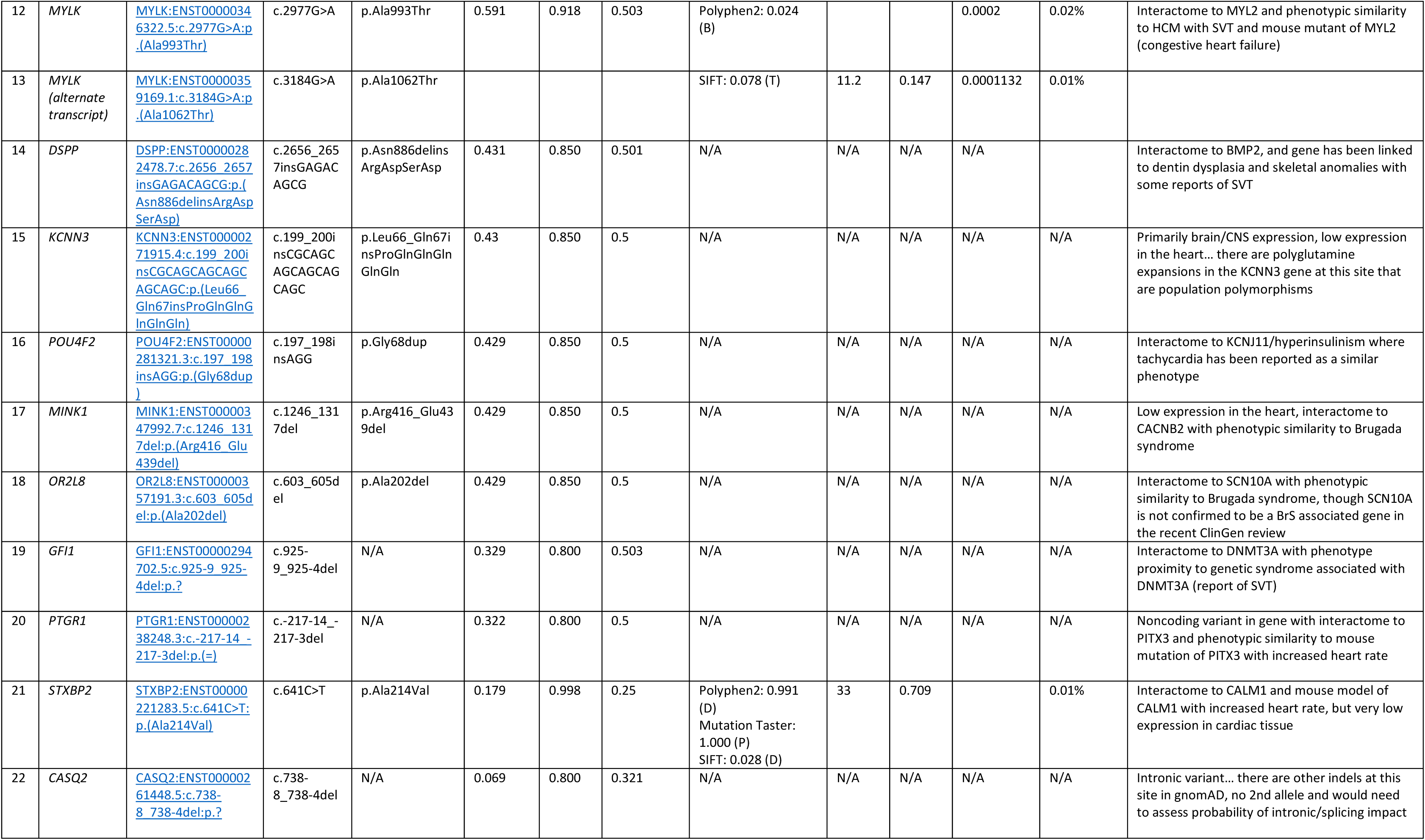
List of all prioritized candidate gene variants with autosomal dominant inheritance and co-segregating with cJET in all affected family members.

## References

1. Villain E, Vetter VL, Garcia JM, Herre J, Cifarelli A, Garson A, Jr. Evolving concepts in the management of congenital junctional ectopic tachycardia. A multicenter study. Circulation. 1990;81:1544–1549. doi: 10.1161/01.cir.81.5.1544

2. Collins KK, Van Hare GF, Kertesz NJ, Law IH, Bar-Cohen Y, Dubin AM, Etheridge SP, Berul CI, Avari JN, Tuzcu V, et al. Pediatric nonpost-operative junctional ectopic tachycardia medical management and interventional therapies. J Am Coll Cardiol. 2009;53:690–697. doi: 10.1016/j.jacc.2008.11.019

3. Dieks JK, Klehs S, Muller MJ, Paul T, Krause U. Adjunctive ivabradine in combination with amiodarone: A novel therapy for pediatric congenital junctional ectopic tachycardia. Heart Rhythm. 2016;13:1297–1302. doi: 10.1016/j.hrthm.2016.03.015

4. Rochelson E, Valdes SO, Asadourian V, Patel R, Lemming K, Howard TS, Pham TDN, Miyake CY, Kim JJ. Sotalol versus amiodarone for postoperative junctional tachycardia after congenital heart surgery. Heart Rhythm. 2022;19:450–456. doi: 10.1016/j.hrthm.2021.11.021

5. Kylat RI, Samson RA. Junctional ectopic tachycardia in infants and children. J Arrhythm. 2020;36:59–66. doi: 10.1002/joa3.12282

6. Borgman KY, Smith AH, Owen JP, Fish FA, Kannankeril PJ. A genetic contribution to risk for postoperative junctional ectopic tachycardia in children undergoing surgery for congenital heart disease. Heart Rhythm. 2011;8:1900–1904. doi: 10.1016/j.hrthm.2011.06.033

7. Milano A, Lodder EM, Bezzina CR. TNNI3K in cardiovascular disease and prospects for therapy. J Mol Cell Cardiol. 2015;82:167–173. doi: 10.1016/j.yjmcc.2015.03.008

8. Xi Y, Honeywell C, Zhang D, Schwartzentruber J, Beaulieu CL, Tetreault M, Hartley T, Marton J, Vidal SM, Majewski J, et al. Whole exome sequencing identifies the TNNI3K gene as a cause of familial conduction system disease and congenital junctional ectopic tachycardia. Int J Cardiol. 2015;185:114–116. doi: 10.1016/j.ijcard.2015.03.130

9. Dumeny L, Chantra M, Langaee T, Duong BQ, Zambrano DH, Han F, Lopez-Colon D, Humma JF, Dacosta J, Lovato T, et al. beta1-receptor polymorphisms and junctional ectopic tachycardia in children after cardiac surgery. Clin Transl Sci. 2022;15:619–625. doi: 10.1111/cts.13178

10. Sarubbi B, Musto B, Ducceschi V, D’Onofrio A, Cavallaro C, Vecchione F, Musto C, Calabro R. Congenital junctional ectopic tachycardia in children and adolescents: a 20 year experience based study. Heart. 2002;88:188–190. doi: 10.1136/heart.88.2.188

11. Pham C, Koopmann TT, Vinocur JM, Blom NA, Nogueira Silbiger V, Mittal K, Bootsma M, Palm KCA, Clur SB, Barge-Schaapveld D, et al. Reduced kinase function in two ultra-rare TNNI3K variants in families with congenital junctional ectopic tachycardia. Clin Genet. 2024;106:37–46. doi: 10.1111/cge.14504

12. Theis JL, Zimmermann MT, Larsen BT, Rybakova IN, Long PA, Evans JM, Middha S, de Andrade M, Moss RL, Wieben ED, et al. TNNI3K mutation in familial syndrome of conduction system disease, atrial tachyarrhythmia and dilated cardiomyopathy. Hum Mol Genet. 2014;23:5793–5804. doi: 10.1093/hmg/ddu297

13. Maiers JA, Ebenroth ES. Junctional ectopic tachycardia following complete heart block associated with viral myocarditis. Pediatr Cardiol. 2006;27:367–368. doi: 10.1007/s00246-005-1215-x

14. Landstrom AP, Yang Q, Sun B, Perelli RM, Bidzimou MT, Zhang Z, Aguilar-Sanchez Y, Alsina KM, Cao S, Reynolds JO, et al. Reduction in Junctophilin 2 Expression in Cardiac Nodal Tissue Results in Intracellular Calcium-Driven Increase in Nodal Cell Automaticity. Circ Arrhythm Electrophysiol. 2023;16:e010858. doi: 10.1161/CIRCEP.122.010858

15. Yang Q, Tadros HJ, Sun B, Bidzimou MT, Ezekian JE, Li F, Ludwig A, Wehrens XHT, Landstrom AP. Junctional Ectopic Tachycardia Caused by Junctophilin-2 Expression Silencing Is Selectively Sensitive to Ryanodine Receptor Blockade. JACC Basic Transl Sci. 2023;8:1577–1588. doi: 10.1016/j.jacbts.2023.07.008

16. Auer PL, Stitziel NO. Genetic association studies in cardiovascular diseases: Do we have enough power? Trends Cardiovasc Med. 2017;27:397–404. doi: 10.1016/j.tcm.2017.03.005

17. Evangelou E, Trikalinos TA, Salanti G, Ioannidis JP. Family-based versus unrelated case-control designs for genetic associations. PLoS Genet. 2006;2:e123. doi: 10.1371/journal.pgen.0020123

18. Wessel J, Schork AJ, Tiwari HK, Schork NJ. Powerful designs for genetic association studies that consider twins and sibling pairs with discordant genotypes. Genet Epidemiol. 2007;31:789–796. doi: 10.1002/gepi.20241

19. Borecki IB, Province MA. Genetic and genomic discovery using family studies. Circulation. 2008;118:1057–1063. doi: 10.1161/CIRCULATIONAHA.107.714592

20. Gruscheski L, Brand T. The Role of POPDC Proteins in Cardiac Pacemaking and Conduction. J Cardiovasc Dev Dis. 2021;8. doi: 10.3390/jcdd8120160

21. Froese A, Breher SS, Waldeyer C, Schindler RF, Nikolaev VO, Rinne S, Wischmeyer E, Schlueter J, Becher J, Simrick S, et al. Popeye domain containing proteins are essential for stress-mediated modulation of cardiac pacemaking in mice. J Clin Invest. 2012;122:1119–1130. doi: 10.1172/JCI59410

22. Rinne S, Kiper AK, Jacob R, Ortiz-Bonnin B, Schindler RFR, Fischer S, Komadowski M, De Martino E, Schafer MK, Cornelius T, et al. Popeye domain containing proteins modulate the voltage-gated cardiac sodium channel Nav1.5. iScience. 2024;27:109696. doi: 10.1016/j.isci.2024.109696

23. Swan AH, Schindler RFR, Savarese M, Mayer I, Rinne S, Bleser F, Schanzer A, Hahn A, Sabatelli M, Perna F, et al. Differential effects of mutations of POPDC proteins on heteromeric interaction and membrane trafficking. Acta Neuropathol Commun. 2023;11:4. doi: 10.1186/s40478-022-01501-w

24. Retterer K, Scuffins J, Schmidt D, Lewis R, Pineda-Alvarez D, Stafford A, Schmidt L, Warren S, Gibellini F, Kondakova A, et al. Assessing copy number from exome sequencing and exome array CGH based on CNV spectrum in a large clinical cohort. Genet Med. 2015;17:623–629. doi: 10.1038/gim.2014.160

25. Richards S, Aziz N, Bale S, Bick D, Das S, Gastier-Foster J, Grody WW, Hegde M, Lyon E, Spector E, et al. Standards and guidelines for the interpretation of sequence variants: a joint consensus recommendation of the American College of Medical Genetics and Genomics and the Association for Molecular Pathology. Genet Med. 2015;17:405–424. doi: 10.1038/gim.2015.30

26. Danecek P, Bonfield JK, Liddle J, Marshall J, Ohan V, Pollard MO, Whitwham A, Keane T, McCarthy SA, Davies RM, et al. Twelve years of SAMtools and BCFtools. Gigascience. 2021;10. doi: 10.1093/gigascience/giab008

27. Bone WP, Washington NL, Buske OJ, Adams DR, Davis J, Draper D, Flynn ED, Girdea M, Godfrey R, Golas G, et al. Computational evaluation of exome sequence data using human and model organism phenotypes improves diagnostic efficiency. Genet Med. 2016;18:608–617. doi: 10.1038/gim.2015.137

28. Cipriani V, Pontikos N, Arno G, Sergouniotis PI, Lenassi E, Thawong P, Danis D, Michaelides M, Webster AR, Moore AT, et al. An Improved Phenotype-Driven Tool for Rare Mendelian Variant Prioritization: Benchmarking Exomiser on Real Patient Whole-Exome Data. Genes (Basel). 2020;11. doi: 10.3390/genes11040460

29. Smedley D, Jacobsen JO, Jager M, Kohler S, Holtgrewe M, Schubach M, Siragusa E, Zemojtel T, Buske OJ, Washington NL, et al. Next-generation diagnostics and disease-gene discovery with the Exomiser. Nat Protoc. 2015;10:2004–2015. doi: 10.1038/nprot.2015.124

30. Pengelly RJ, Alom T, Zhang Z, Hunt D, Ennis S, Collins A. Evaluating phenotype-driven approaches for genetic diagnoses from exomes in a clinical setting. Sci Rep. 2017;7:13509. doi: 10.1038/s41598-017-13841-y

31. Jacobsen JOB, Kelly C, Cipriani V, Robinson PN, Smedley D. Evaluation of phenotype-driven gene prioritization methods for Mendelian diseases. Brief Bioinform. 2022;23. doi: 10.1093/bib/bbac188

32. Ioannidis NM, Rothstein JH, Pejaver V, Middha S, McDonnell SK, Baheti S, Musolf A, Li Q, Holzinger E, Karyadi D, et al. REVEL: An Ensemble Method for Predicting the Pathogenicity of Rare Missense Variants. Am J Hum Genet. 2016;99:877–885. doi: 10.1016/j.ajhg.2016.08.016

33. Kircher M, Witten DM, Jain P, O’Roak BJ, Cooper GM, Shendure J. A general framework for estimating the relative pathogenicity of human genetic variants. Nat Genet. 2014;46:310–315. doi: 10.1038/ng.2892

34. Niroula A, Vihinen M. How good are pathogenicity predictors in detecting benign variants? PLoS Comput Biol. 2019;15:e1006481. doi: 10.1371/journal.pcbi.1006481

35. Schindelin J, Arganda-Carreras I, Frise E, Kaynig V, Longair M, Pietzsch T, Preibisch S, Rueden C, Saalfeld S, Schmid B, et al. Fiji: an open-source platform for biological-image analysis. Nat Methods. 2012;9:676–682. doi: 10.1038/nmeth.2019

36. Jumper J, Evans R, Pritzel A, Green T, Figurnov M, Ronneberger O, Tunyasuvunakool K, Bates R, Zidek A, Potapenko A, et al. Highly accurate protein structure prediction with AlphaFold. Nature. 2021;596:583–589. doi: 10.1038/s41586-021-03819-2

37. Pettersen EF, Goddard TD, Huang CC, Couch GS, Greenblatt DM, Meng EC, Ferrin TE. UCSF Chimera--a visualization system for exploratory research and analysis. J Comput Chem. 2004;25:1605–1612. doi: 10.1002/jcc.20084

38. Krieger E, Vriend G. New ways to boost molecular dynamics simulations. J Comput Chem. 2015;36:996–1007. doi: 10.1002/jcc.23899

39. Krieger E, Dunbrack RL, Jr., Hooft RW, Krieger B. Assignment of protonation states in proteins and ligands: combining pKa prediction with hydrogen bonding network optimization. Methods Mol Biol. 2012;819:405–421. doi: 10.1007/978-1-61779-465-0_25

40. Maier JA, Martinez C, Kasavajhala K, Wickstrom L, Hauser KE, Simmerling C. ff14SB: Improving the Accuracy of Protein Side Chain and Backbone Parameters from ff99SB. J Chem Theory Comput. 2015;11:3696–3713. doi: 10.1021/acs.jctc.5b00255

41. Hornak V, Abel R, Okur A, Strockbine B, Roitberg A, Simmerling C. Comparison of multiple Amber force fields and development of improved protein backbone parameters. Proteins. 2006;65:712–725. doi: 10.1002/prot.21123

42. Essmann U, Perera L, Berkowitz ML, Darden T, Lee H, Pedersen LG. A smooth particle mesh Ewald method. J Chem Phys. 1995;103:8577–8593.

43. Schindler RF, Scotton C, Zhang J, Passarelli C, Ortiz-Bonnin B, Simrick S, Schwerte T, Poon KL, Fang M, Rinne S, et al. POPDC1(S201F) causes muscular dystrophy and arrhythmia by affecting protein trafficking. J Clin Invest. 2016;126:239–253. doi: 10.1172/JCI79562

44. Kirchmaier BC, Poon KL, Schwerte T, Huisken J, Winkler C, Jungblut B, Stainier DY, Brand T. The Popeye domain containing 2 (popdc2) gene in zebrafish is required for heart and skeletal muscle development. Dev Biol. 2012;363:438–450. doi: 10.1016/j.ydbio.2012.01.015

45. Rinne S, Ortiz-Bonnin B, Stallmeyer B, Kiper AK, Fortmuller L, Schindler RFR, Herbort-Brand U, Kabir NS, Dittmann S, Friedrich C, et al. POPDC2 a novel susceptibility gene for conduction disorders. J Mol Cell Cardiol. 2020;145:74–83. doi: 10.1016/j.yjmcc.2020.06.005

46. Baldwin TA, Li Y, Marsden AN, Rinne S, Garza-Carbajal A, Schindler RFR, Zhang M, Garcia MA, Venna VR, Decher N, et al. POPDC1 scaffolds a complex of adenylyl cyclase 9 and the potassium channel TREK-1 in heart. EMBO Rep. 2022;23:e55208. doi: 10.15252/embr.202255208

47. Tibbo AJ, Mika D, Dobi S, Ling J, McFall A, Tejeda GS, Blair C, MacLeod R, MacQuaide N, Gok C, et al. Phosphodiesterase type 4 anchoring regulates cAMP signaling to Popeye domain-containing proteins. J Mol Cell Cardiol. 2022;165:86–102. doi: 10.1016/j.yjmcc.2022.01.001

48. Creighton TE. Proteins: structures and molecular properties. 2nd ed. W.H. Freeman; 1993.

49. Podliesna S, Delanne J, Miller L, Tester DJ, Uzunyan M, Yano S, Klerk M, Cannon BC, Khongphatthanayothin A, Laurent G, et al. Supraventricular tachycardias, conduction disease, and cardiomyopathy in 3 families with the same rare variant in TNNI3K (p.Glu768Lys). Heart Rhythm. 2019;16:98–105. doi: 10.1016/j.hrthm.2018.07.015

50. Pham C, Munoz-Martin N, Lodder EM. The Diverse Roles of TNNI3K in Cardiac Disease and Potential for Treatment. Int J Mol Sci. 2021;22. doi: 10.3390/ijms22126422

51. Brand T, Simrick SL, Poon KL, Schindler RF. The cAMP-binding Popdc proteins have a redundant function in the heart. Biochem Soc Trans. 2014;42:295–301. doi: 10.1042/BST20130264

52. Schindler RF, Brand T. The Popeye domain containing protein family--A novel class of cAMP effectors with important functions in multiple tissues. Prog Biophys Mol Biol. 2016;120:28–36. doi: 10.1016/j.pbiomolbio.2016.01.001

53. Beecher G, Tang C, Liewluck T. Severe adolescent-onset limb-girdle muscular dystrophy due to a novel homozygous nonsense BVES variant. J Neurol Sci. 2021;420:117259. doi: 10.1016/j.jns.2020.117259

54. De Ridder W, Nelson I, Asselbergh B, De Paepe B, Beuvin M, Ben Yaou R, Masson C, Boland A, Deleuze JF, Maisonobe T, et al. Muscular dystrophy with arrhythmia caused by loss-of-function mutations in BVES. Neurol Genet. 2019;5:e321. doi: 10.1212/NXG.0000000000000321

55. Gangfuss A, Hentschel A, Heil L, Gonzalez M, Schonecker A, Depienne C, Nishimura A, Zengeler D, Kohlschmidt N, Sickmann A, et al. Proteomic and morphological insights and clinical presentation of two young patients with novel mutations of BVES (POPDC1). Mol Genet Metab. 2022;136:226–237. doi: 10.1016/j.ymgme.2022.05.005

56. Mahmood A, Samad A, Shah AA, Wadood A, Alkathiri A, Alshehri MA, Alam MZ, Hussain T, He P, Umair M. A novel biallelic variant in the Popeye domain-containing protein 1 (POPDC1) underlies limb girdle muscle dystrophy type 25. Clin Genet. 2023;103:219–225. doi: 10.1111/cge.14238

57. Indrawati LA, Iida A, Tanaka Y, Honma Y, Mizoguchi K, Yamaguchi T, Ikawa M, Hayashi S, Noguchi S, Nishino I. Two Japanese LGMDR25 patients with a biallelic recurrent nonsense variant of BVES. Neuromuscul Disord. 2020;30:674–679. doi: 10.1016/j.nmd.2020.06.004

58. Andree B, Hillemann T, Kessler-Icekson G, Schmitt-John T, Jockusch H, Arnold HH, Brand T. Isolation and characterization of the novel popeye gene family expressed in skeletal muscle and heart. Dev Biol. 2000;223:371–382. doi: 10.1006/dbio.2000.9751

59. Hamilton S, Terentyev D. ER stress and calcium-dependent arrhythmias. Front Physiol. 2022;13:1041940. doi: 10.3389/fphys.2022.1041940

60. Ciccone MP, Panfili FM, Bacigalupo F, Brusco F, Florean L, Re F, Bottillo I. A Novel POPDC2 Pathogenic Variant in a Young Patient With Cardiac Conduction Disease and Hypertrophic Cardiomyopathy. Clin Genet. 2026;109:986–988. doi: 10.1111/cge.70131

61. Nicastro M, Vermeer AMC, Postema PG, Tadros R, Bowling FZ, Aegisdottir HM, Tragante V, Mach L, Postma AV, Lodder EM, et al. Bi-allelic variants in POPDC2 cause an autosomal recessive syndrome presenting with cardiac conduction defects and hypertrophic cardiomyopathy. Am J Hum Genet. 2025;112:1681–1698. doi: 10.1016/j.ajhg.2025.04.016

62. Li H, Wang P, Zhang C, Zuo Y, Zhou Y, Han R. Defective BVES-mediated feedback control of cAMP in muscular dystrophy. Nat Commun. 2023;14:1785. doi: 10.1038/s41467-023-37496-8

63. Mulkey SB, Plessis AD. The Critical Role of the Central Autonomic Nervous System in Fetal-Neonatal Transition. Semin Pediatr Neurol. 2018;28:29–37. doi: 10.1016/j.spen.2018.05.004

